# Predictors of uncertainty and unwillingness to receive the COVID-19 booster vaccine: an observational study of 22,139 fully vaccinated adults in the UK

**DOI:** 10.1101/2021.12.17.21267941

**Authors:** Elise Paul, Daisy Fancourt

**Author notes:** Corresponding author: Dr Elise Paul, 1-19 Torrington Place, London, WC1E 7HB, United Kingdom.

## Abstract

**Background:** The continued success of the COVID-19 vaccination programme in the UK will depend on widespread uptake of booster vaccines. However, there is evidence of hesitancy and unwillingness to receive the booster vaccine, even in fully vaccinated adults. Identifying factors associated with COVID-19 booster vaccine intentions specifically in this population is therefore critical.

**Methods:** We used data from 22,139 fully vaccinated adults who took part in the UCL COVID-19 Social Study. Multinomial logistic regression examined predictors of uncertainty and unwillingness (versus willingness) to receive a COVID-19 booster vaccine (measured 22 November 2021 to 6 December 2021), including (i) socio-demographic factors, (ii) COVID-19 related factors (e.g., having been infected with COVID-19), and (iii) initial intent to receive a COVID-19 vaccine in the four months following the announcement in the UK that the vaccines had been approved (2 December 2020 to 31 March 2021).

**Findings:** 4% of the sample reported that they were uncertain about receiving a COVID-19 booster vaccine, and a further 4% unwilling. Initial uncertainty and unwillingness to accept the first COVID-19 vaccine in 2020-21 were each associated with over five times the risk of being uncertain about and unwilling to accept a booster vaccine. Healthy adults (those without a pre-existing physical health condition) were also more likely to be uncertain or unwilling to receive a booster vaccine. In addition, low levels of current stress about catching or becoming seriously ill from COVID-19, consistently low compliance with COVID-19 government guidelines during periods of strict restrictions (e.g., lockdowns), lower levels of educational qualification, lower socio-economic position, and age below 45 years were all associated with uncertainty and unwillingness.

**Interpretation:** Our findings highlight that there are a range of factors that predict booster intentions, with the strongest predictor being previous uncertainty and unwillingness. Two other concerning patterns also emerged from our results. First, administration of booster vaccinations may increase social inequalities in experiences of COVID-19 as adults from lower socio-economic backgrounds are also most likely to be uncertain or unwilling to accept a booster vaccine as well as most likely to be seriously affected by the virus. Second, some of those most likely to spread COVID-19 (i.e., those with poor compliance with guidelines) are most likely to be uncertain and unwilling. Public health messaging should be tailored specifically to these groups.

**Funding:** The Nuffield Foundation [WEL/FR-000022583], the MARCH Mental Health Network funded by the Cross-Disciplinary Mental Health Network Plus initiative supported by UK Research and Innovation [ES/S002588/1], and the Wellcome Trust [221400/Z/20/Z and 205407/Z/16/Z].

## Introduction

Although highly effective in preventing severe disease and death,^1,2^ emerging evidence suggests that immunity from COVID-19 vaccines diminishes over time.^3,4^ Licensed COVID-19 vaccines may also be less effective against new more transmissible variants^5,6^ and may therefore need to be updated.^7^ Consequently, in response to the new Omicron wave of the virus in the autumn of 2021, some countries, such as the UK, announced plans to introduce booster vaccinations. This announcement was not without controversy, particularly given the continued unequal access to vaccinations worldwide.^8^ Nonetheless, the strategic rationale behind this decision was that widespread uptake of COVID-19 booster vaccines in the general population could help to prevent further deaths and serious illness due to COVID-19 and reduce the burden on the healthcare system whilst reducing the need for more stringent social restrictions.

Although COVID-19 vaccine uptake in the UK has, overall, been high,^9^ some groups have been more likely than others to refuse the vaccine.^10^ People from ethnic minority groups, those who have already been infected with COVID-19, people living in more deprived areas, people who are unemployed or not working, and smokers are less likely to have received at least one dose of a COVID-19 vaccine.^10^ Some of these same groups had also expressed hesitancy or unwillingness to receive a COVID-19 vaccine before they were available: people with lower levels of education,^11–16^ lower household incomes,^11,12,17^ and people from ethnic minority groups.^13,15,16^ However, other demographic groups reporting COVID-19 vaccine hesitancy before the vaccines were available^11^ have not shown differential uptake in the UK (e.g., women).^10^

Recent data from the Opinions and Lifestyle Survey (OPN) in the UK suggest a considerable proportion of the general population who have received two doses of a COVID-19 vaccine are either unwilling (5%; very unlikely or fairly unlikely) or unsure (5%; neither likely nor unlikely) about accepting a booster vaccine.^18^ Young adults (ages 16-29; 9%) and adults over the age of 70 (10%) were more than twice as likely as other age groups (3% of adults ages 30-49 and 1% of adults ages 50-69) to say they are unsure about accepting a booster, whilst the proportion of men saying they would refuse was nearly twice that of women (6% vs 4%).^19^ The proportion of fully vaccinated adults who previously reported they were unlikely or unsure about having a first dose of a COVID-19 vaccine is even higher (22%).^20^ But further research into predictors of booster hesitancy or unwillingness is lacking.

Therefore, the aims of this study were to identify associations between socio-demographic and COVID-19 related factors (e.g., having been infected with COVID-19) and initial intent to receive the COVID-19 vaccine following the announcement in the UK that the vaccines had been approved (2 December 2020 to 31 March 2021) with uncertainty and unwillingness to receive a COVID-19 booster vaccine in adults who had been fully vaccinated.

## Methods

### Study design and participants

Data were drawn from the COVID-19 Social Study; a large population-based panel study of the psychological (e.g., symptoms of depression and anxiety, worry about COVID-19 related adversities) and social experiences (e.g., loneliness, psychological and physical abuse) of over 70,000 adults (aged 18+) in the UK during the COVID-19 pandemic. See the study User Guide for a complete listing of study questions and modules (https://osf.io/jm8ra/). The study commenced on 21 March 2020 and involves online weekly data collection from participants for the duration of the COVID-19 pandemic in the UK. The study is not random and therefore is not representative of the UK population. But it does contain a well-stratified sample that was recruited using three primary approaches. First, convenience sampling was used, including promoting the study through existing networks and mailing lists (including large databases of adults who had previously consented to be involved in health research across the UK), print and digital media coverage, and social media. Second, more targeted recruitment was undertaken focusing on groups who were anticipated to be less likely to take part in the research via our first strategy, including (i) individuals from a low-income background, (ii) individuals with no or few educational qualifications, and (iii) individuals who were unemployed. Third, the study was promoted via partnerships with third sector organisations to vulnerable groups, including adults with pre-existing mental health conditions, older adults, carers, and people experiencing domestic violence or abuse. The study was approved by the UCL Research Ethics Committee [12467/005] and all participants gave informed consent. Participants were not compensated for participation.

In the current study, we included individuals who had (i) data on intent to receive a COVID-19 booster vaccine (collected 22 November to 6 December 2021, hereafter ‘follow-up’), (ii) data on initial intent to receive a first COVID-19 vaccine (collected 2 December 2020 to 31 March 2021), (iii) received at least two doses of a COVID-19 vaccine at follow-up, and (iv) complete data on other study variables.

A total of 32,096 participants met the first criterion, and of these, 26,619 also had data on initial COVID-19 vaccine intent, and 25,821 also met the third criterion of having had at least two doses of a COVID-19 vaccine at follow-up. 3,682 individuals were excluded due to missing data on other study variables, for example those who had selected “other/prefer not to say” in response to gender or “prefer not to say” on ethnicity or income, due to insufficient statistical power and an inability to create statistical weights for these individuals, leaving a total analytical sample of 22,139. See Supplemental Table S1 for a comparison of excluded and included participants on study variables.

## Measures

### Outcome variable

On 14 September 2021, the UK Joint Committee on vaccination and Immunisation (JCVI) announced that booster vaccinations were eligible to vulnerable groups (adults over the age of 50, health and social are workers and people who were clinically vulnerable) providing they had had their second COVID vaccination at least six months ago. On 29 November 2021 in response to the rise of the Omicron variant, the UK government accelerated the booster programme by announcing that all adults over the age of 18 would be eligible to receive a booster vaccine at least three months after their second dose.

Intention to receive a COVID-19 booster vaccine was measured in the November COVID-19 Social Study with a study developed item (“How likely do you think you are to get a COVID-19 booster vaccine if/when you are offered one?”). Response options ranged from 1 “very unlikely” to 6 “very likely”, as well as 7 “I have already had one/accepted one”. An ordinal variable was coded: (0) intend to receive the booster vaccine (responses of 5-7), (1) unsure about whether to receive the booster vaccine (responses of 3-4), and (2) unwilling to receive the booster vaccine (responses of 1-2). Participants who had already received or accepted a booster vaccine were included in the comparison group in order to maximise sample size and ensure that older and vulnerable groups were part of analyses, under the assumption that uptake of the booster vaccine so early in the programme indicated a willingness to receive it. To examine how their inclusion may have affected results, a sensitivity analysis which excluded these people was carried out. See Supplemental Table S2 for a listing of all question wording for study developed items.

### Predictor variables

#### Socio-demographics

Socio-demographic factors were collected at baseline (i.e., each individual’s first participation in the study [21 March to 6 September 2020], which occurred before the measurement of other study variables) and included gender (male vs female), age group (60+, 45-59, 30-44, and 18-29) ethnicity (white vs ethnic minority groups [i.e., Asian/Asian British, Black/Black British. See Supplemental Table S2 for a full listing of response options]), education (undergraduate degree or higher [further education after the age of 18], A-levels (equivalent to education to age 18) or vocational training, and up to GCSE (General Certificate of Secondary Education) [equivalent to education to age 16]), income (annual household income: >£90,0000, £60,000-89,999, £30,000-59,999, £16,000-29,999, and < £16,000), smoking status (non-smoker vs former or current smoker), employment status (not employed [still at school, at university, unable to work due to disability, unemployed and seeking work, or retired vs employed [self-employed, part-time or full-time employed]), area of dwelling (urban [city, large town, small town] vs rural [village, hamlet, isolated dwelling]), living arrangement (living alone vs living with others but not including children vs living with others, including children), and government’s identified key worker status (not a key worker vs key worker). The latter included people with jobs deemed essential during the pandemic (e.g., health and social care, education, and childcare) and who were required to leave home to carry out this work during lockdowns.

Participants reported at baseline whether they had received a clinical diagnosis of a mental health condition (e.g., depression, anxiety, or other psychiatric diagnosis) or chronic physical health condition (e.g., high blood pressure, diabetes, or other physical health condition) were used to create two binary variables (yes/no) to indicate the presence or absence of pre-existing physical and mental health conditions.

### COVID-19 related factors

Initial intent to receive a COVID-19 vaccine was measured with a study developed item administered 2 December 2020 to 31 March 2021. Many participants had vaccine intention data for all four months during this period, but because we were interested in their first recorded intention to receive the vaccine after its authorisation was announced in the UK on 2 December 2020, we coded initial intent in this way. In a sensitivity analysis, participants’ last recorded COVID-19 vaccine intention during the period 2 December 2020 to 31 March 2021 was used.

Ever having had COVID-19 was operationalised from a study developed item administered at follow-up which was coded as: yes, confirmed by a test (either a positive COVID-19 test or an antibody test), yes, suspected by either a doctor or the participant, and no.

Confidence in government and the health service to handle the pandemic were assessed with one study developed question each at follow-up. Response options ranged from 1 (none at all) to 7 (lots). Two binary variables were created to indicate a lot of (5-7) versus low (1-4) confidence in the government and health system.

Self-reported compliance with government COVID-19 guidelines was measured with one study developed question with response options ranging from 1 (none at all) to 7 (very much so). We analyse this as an binary variable reflecting patterns of compliance whilst restrictions were in place across the pandemic. As most of our sample resides in England, we used dates for which there were strict COVID-19 restrictions in England (e.g., rather than Scotland or Northern Ireland). See Supplemental Table S3 for a listing of these dates. The binary variable was coded as: consistently high compliance (a mean score of 6-7 across all strict restriction periods) vs consistently low compliance (mean score of 1-5 across all strict restriction periods). Sensitivity analyses using i) only participants with compliance data from all three strict restriction periods and ii) a variable reflecting compliance across the entirety of the pandemic were conducted.

Self-reported knowledge of COVID-19 was measured at follow-up with one study developed question and rated on a 7-point scale from 1 (very poor knowledge) to 7 (very good knowledge). Responses of 1-4 were categorised as low compared to high (5-7) COVID-19 knowledge. The presence or absence of worry about either contracting COVID-19 or becoming seriously ill from it were captured from two multiple choice questions asked at follow-up. A binary variable was created to indicate not having endorsed either as a source of stress vs having endorsed one or both.

### Statistical analysis

Two multinomial regression models were fitted to examine associations between socio-demographic and COVID-19 related factors and uncertainty and unwillingness to receive a COVID-19 booster vaccine. The first model included only socio-demographic variables whilst the second additionally included COVID-19 related variables. The outcome variable in these two models was coded such that those who were uncertain or unwilling were compared to those who were very likely to receive the COVID-19 booster vaccine or had already accepted one. Multinomial regression coefficients were exponentiated and presented as relative risk ratios (RRR) with corresponding 95% confidence intervals (CI).

To account for the non-random nature of the sample and increase representativeness of the UK general population, all data were weighted to the proportions of gender, age, ethnicity, country, and education obtained from the Office for National Statistics.^21^ A multivariate reweighting method was implemented using the Stata user written command ‘ebalance’.^22^ Analyses were conducted using Stata version 16.^23^

## Results

Unweighted and weighted sample characteristics are presented in Table 1. Before weighting, the sample was disproportionately female, older (ages 60+), white, and had education levels of at least an undergraduate degree. But after weighting the sample aligned with expected population characteristics. Figure 1 presents proportions of the weighted sample expressing willingness, uncertainty, and unwillingness to receive the booster vaccine by initial COVID-19 vaccine intent from a cross-tabulation. One in five (19· 8%) adults who were initially unwilling to receive a vaccine but did nonetheless have two vaccinations were unwilling to accept a booster. A further 12· 2% who were uncertain about receiving a first dose of a COVID-19 vaccine (but still went on to have two vaccinations) were unwilling to accept a booster. This is compared to just 2· 4% of those who were initially willing to be vaccinated.

**Table 1.**
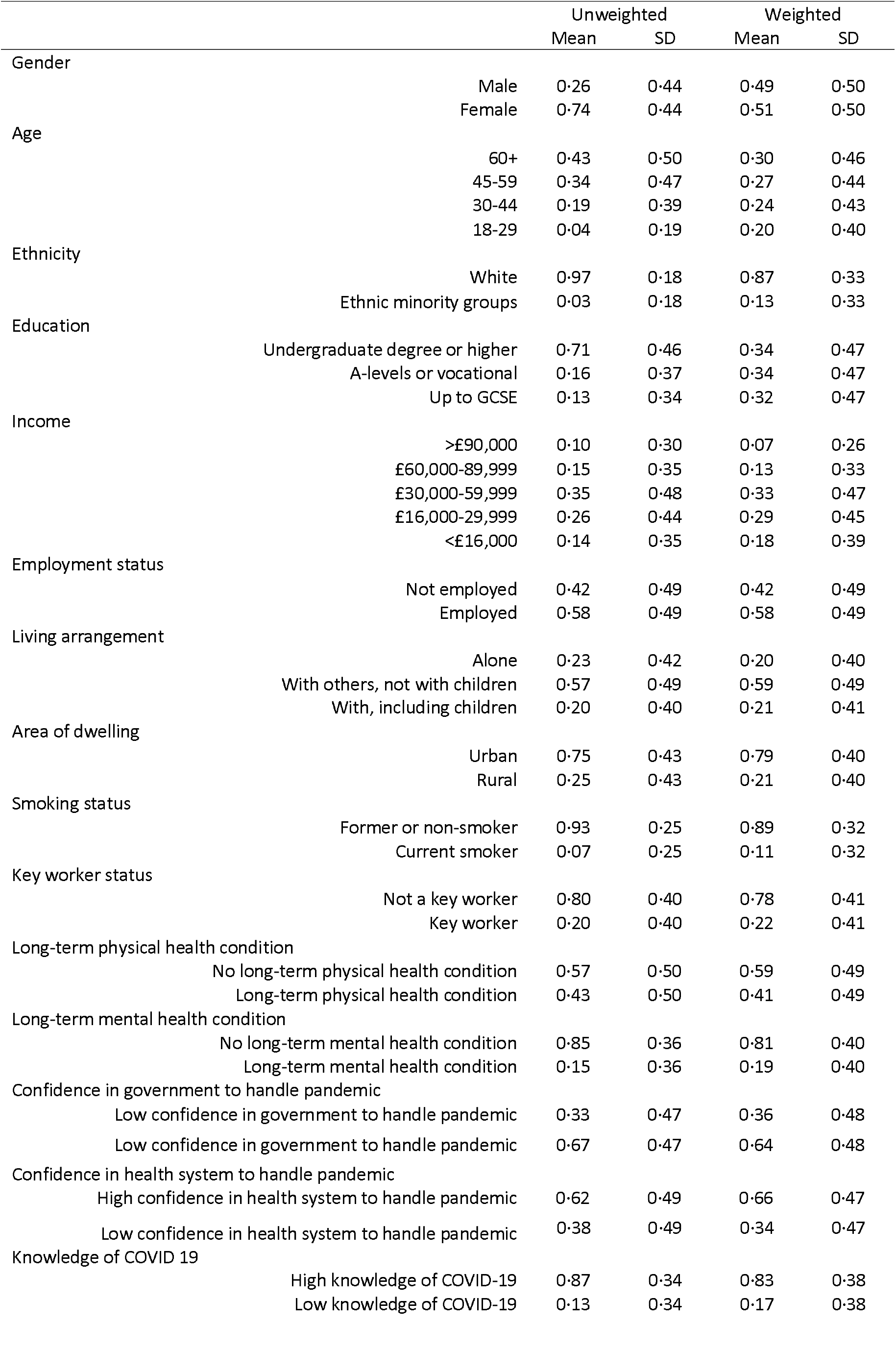
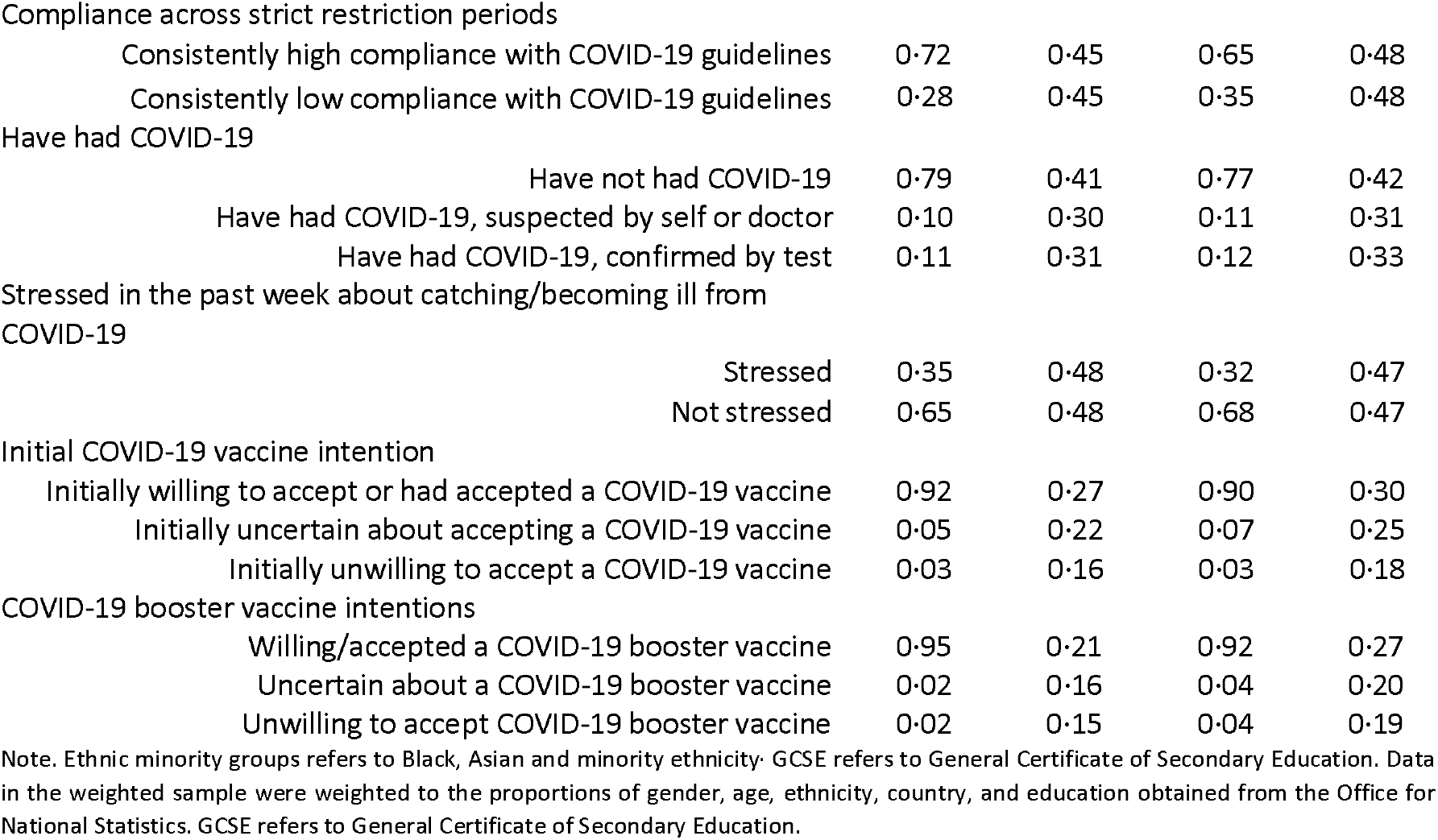
Unweighted and weighted sample characteristics (*N* = 22,139)

|  |  | Unweighted |  | Weighted |  |
| --- | --- | --- | --- | --- | --- |
|  |  | Mean | SD | Mean | SD |
| Gender | Male | 0.26 | 0.44 | 0.49 | 0.50 |
|  | Female | 0.74 | 0.44 | 0.51 | 0.50 |
| Age | 60+ | 0.43 | 0.50 | 0.30 | 0.46 |
|  | 45-59 | 0.34 | 0.47 | 0.27 | 0.44 |
|  | 30-44 | 0.19 | 0.39 | 0.24 | 0.43 |
|  | 18-29 | 0.04 | 0.19 | 0.20 | 0.40 |
| Ethnicity | White | 0.97 | 0.18 | 0.87 | 0.33 |
|  | Ethnic minority groups | 0.03 | 0.18 | 0.13 | 0.33 |
| Education | Undergraduate degree or higher | 0.71 | 0.46 | 0.34 | 0.47 |
|  | A-levels or vocational | 0.16 | 0.37 | 0.34 | 0.47 |
|  | Up to GCSE | 0.13 | 0.34 | 0.32 | 0.47 |
| Income | >£90,000 | 0.10 | 0.30 | 0.07 | 0.26 |
|  | £60,000-89,999 | 0.15 | 0.35 | 0.13 | 0.33 |
|  | £30,000-59,999 | 0.35 | 0.48 | 0.33 | 0.47 |
|  | £16,000-29,999 | 0.26 | 0.44 | 0.29 | 0.45 |
|  | <£16,000 | 0.14 | 0.35 | 0.18 | 0.39 |
| Employment status | Not employed | 0.42 | 0.49 | 0.42 | 0.49 |
|  | Employed | 0.58 | 0.49 | 0.58 | 0.49 |
| Living arrangement | Alone | 0.23 | 0.42 | 0.20 | 0.40 |
|  | With others, not with children | 0.57 | 0.49 | 0.59 | 0.49 |
|  | With, including children | 0.20 | 0.40 | 0.21 | 0.41 |
| Area of dwelling | Urban | 0.75 | 0.43 | 0.79 | 0.40 |
|  | Rural | 0.25 | 0.43 | 0.21 | 0.40 |
| Smoking status | Former or non-smoker | 0.93 | 0.25 | 0.89 | 0.32 |
|  | Current smoker | 0.07 | 0.25 | 0.11 | 0.32 |
| Key worker status | Not a key worker | 0.80 | 0.40 | 0.78 | 0.41 |
|  | Key worker | 0.20 | 0.40 | 0.22 | 0.41 |
| Long-term physical health condition | No long-term physical health condition | 0.57 | 0.50 | 0.59 | 0.49 |
|  | Long-term physical health condition | 0.43 | 0.50 | 0.41 | 0.49 |
| Long-term mental health condition | No long-term mental health condition | 0.85 | 0.36 | 0.81 | 0.40 |
|  | Long-term mental health condition | 0.15 | 0.36 | 0.19 | 0.40 |
| Confidence in government to handle pandemic | Low confidence in government to handle pandemic | 0.33 | 0.47 | 0.36 | 0.48 |
|  | High confidence in government to handle pandemic | 0.67 | 0.47 | 0.64 | 0.48 |
| Confidence in health system to handle pandemic | High confidence in health system to handle pandemic | 0.62 | 0.49 | 0.66 | 0.47 |
|  | Low confidence in health system to handle pandemic | 0.38 | 0.49 | 0.34 | 0.47 |
| Knowledge of COVID 19 | High knowledge of COVID-19 | 0.87 | 0.34 | 0.83 | 0.38 |
|  | Low knowledge of COVID-19 | 0.13 | 0.34 | 0.17 | 0.38 |

|  |  |  |  |  |
| --- | --- | --- | --- | --- |
| Compliance across strict restriction periods |  |  |  |  |
| Consistently high compliance with COVID-19 guidelines | 0.72 | 0.45 | 0.65 | 0.48 |
| Consistently low compliance with COVID-19 guidelines | 0.28 | 0.45 | 0.35 | 0.48 |
| Have had COVID-19 |  |  |  |  |
| Have not had COVID-19 | 0.79 | 0.41 | 0.77 | 0.42 |
| Have had COVID-19, suspected by self or doctor | 0.10 | 0.30 | 0.11 | 0.31 |
| Have had COVID-19, confirmed by test | 0.11 | 0.31 | 0.12 | 0.33 |
| Stressed in the past week about catching/becoming ill from COVID-19 |  |  |  |  |
| Stressed | 0.35 | 0.48 | 0.32 | 0.47 |
| Not stressed | 0.65 | 0.48 | 0.68 | 0.47 |
| Initial COVID-19 vaccine intention |  |  |  |  |
| Initially willing to accept or had accepted a COVID-19 vaccine | 0.92 | 0.27 | 0.90 | 0.30 |
| Initially uncertain about accepting a COVID-19 vaccine | 0.05 | 0.22 | 0.07 | 0.25 |
| Initially unwilling to accept a COVID-19 vaccine | 0.03 | 0.16 | 0.03 | 0.18 |
| COVID-19 booster vaccine intentions |  |  |  |  |
| Willing/accepted a COVID-19 booster vaccine | 0.95 | 0.21 | 0.92 | 0.27 |
| Uncertain about a COVID-19 booster vaccine | 0.02 | 0.16 | 0.04 | 0.20 |
| Unwilling to accept COVID-19 booster vaccine | 0.02 | 0.15 | 0.04 | 0.19 |

Note. Ethnic minority groups refers to Black, Asian and minority ethnicity. GCSE refers to General Certificate of Secondary Education. Data in the weighted sample were weighted to the proportions of gender, age, ethnicity, country, and education obtained from the Office for National Statistics. GCSE refers to General Certificate of Secondary Education.

**Figure 1.**
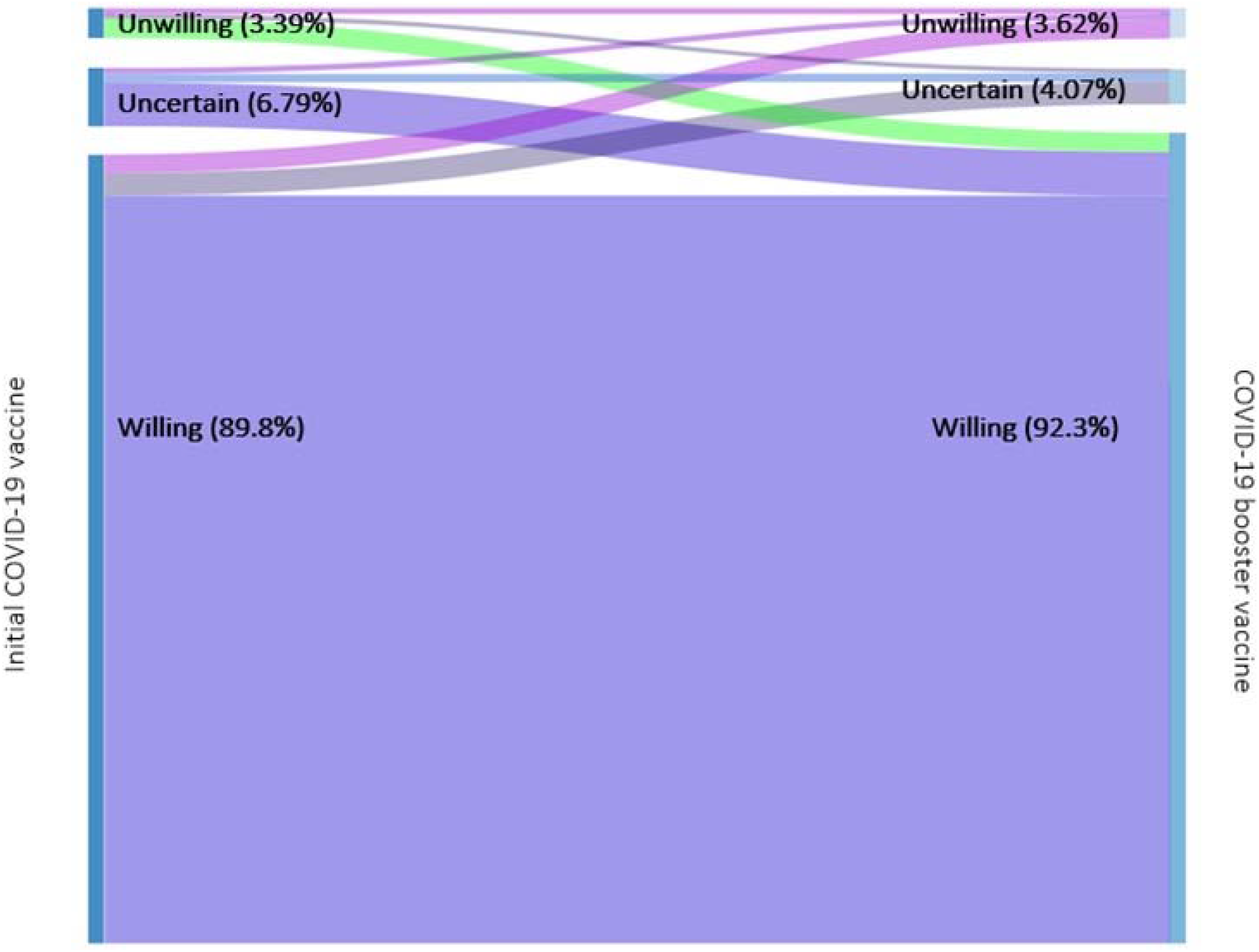
Changes in willingness to receive a COVID-19 vaccine (2 December 2020 to 31 March) and a COVID-19 booster vaccine (22 November to 6 December 2021) (weighted, *N* = 22,139)

4% of the weighted sample expressed unwillingness to receive a COVID-19 booster vaccine (e.g., a score of 1-2 on a scale of 1 to 6), whilst 4% were uncertain (a score of 3-4) (Figure 1). Descriptive differences between adults who were unwilling or uncertain about receiving a COVID-19 booster vaccine vs those who were willing by several demographic characteristics are shown in Table S4.

Of those who were initially willing to accept a COVID-19 vaccine when it became available, 5% said they were either uncertain (3%) or unwilling (2%) to accept a booster (Table 2). 73% of those who were initially uncertain and 65% of those who were initially unwilling had become willing to accept a booster. But 12% of adults who were initially uncertain about accepting a COVID-19 vaccine were now unwilling to accept a booster, and 1 in 5 (20%) who were initially unwilling about a first dose remained unwilling to accept a booster vaccine.

**Table 2.** Proportion of adults in each COVID-19 booster vaccine intent category by initial COVID-19 vaccine intention (weighted, *N* = 22,319)

|  | Initially willing to<br>accept a COVID-19<br>vaccine | Initially uncertain<br>about accepting a<br>COVID-19 vaccine | Initially unwilling<br>to accept a<br>COVID-19<br>vaccine |
| --- | --- | --- | --- |
|  | Prop. | Prop. | Prop. |
| COVID-19 booster vaccine intention |  |  |  |
| Willing to accept or have accepted a booster | 94·77% | 73·48% | 64·80% |
| Uncertain about accepting a booster | 2·87% | 14·37% | 15·39% |
| Unwilling to accept a COVID-19 booster | 2·37% | 12·15% | 19·81% |

Note. Data were weighted to the proportions of gender, age, ethnicity, country, and education obtained from the Office for National Statistics.

### Socio-demographic predictors of booster uncertainty and unwillingness

Results from the multinomial regression model predicting uncertainty and unwillingness to receive a COVID-19 booster vaccine from socio-demographic factors are shown in Table 3. The strongest predictor of booster unwillingness was younger age. Young adults were nearly six times as likely than older adults to be unwilling to receive a booster (RRR = 5· 74; 95% CI: 2· 82 to 11· 68), and adults ages 30-44 were more than twice as likely (RRR = 2· 56; 95% CI: 1· 31 to 5· 00). Education to A-levels or vocational and up to a GCSE were each associated with a 2· 1-2· 5 times higher relative risk of being unwilling to receive a COVID-19 booster vaccine relative to people who held to degrees (A-levels or vocational: RRR = 2· 09; 95% CI: 1· 15 to 3· 79; up to GCSE: RRR = 2· 50; 95% CI: 1· 31 to 4· 79). Additionally, healthy adults were also at increased risk of being unwilling (RRR 1· 52; 95% CI: 1· 00 to 2· 30) to receive the booster compared to individuals with long-term physical health conditions. Ethnicity, living arrangement, smoking status, keyworker status, employment status, income, and having a mental health condition were not associated with uncertainty or unwillingness.

**Table 3.**
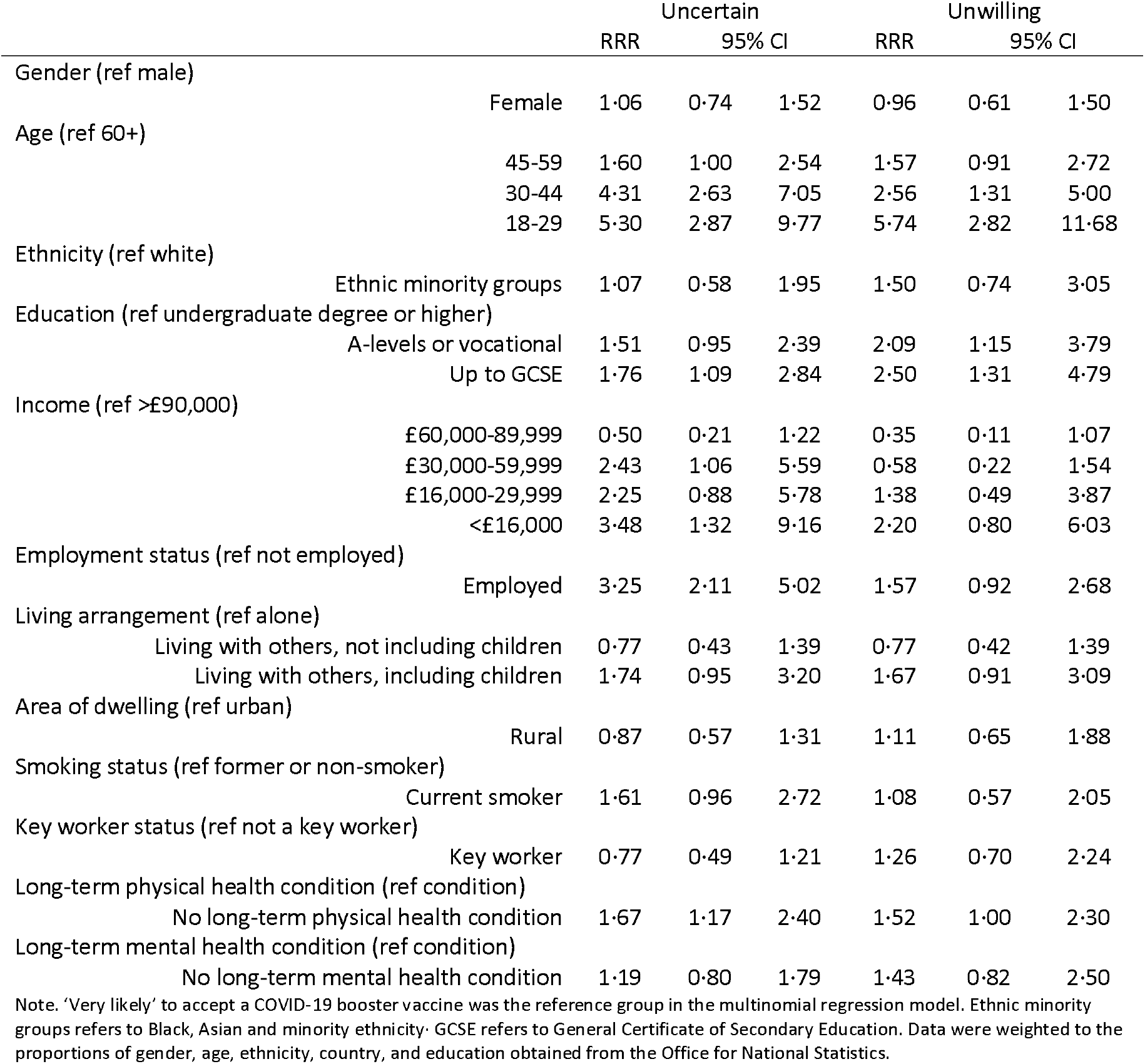
Socio-demographic predictors of uncertainty and unwillingness to receive a COVID-19 booster vaccine using a multivariable multinomial regression (weighted, *N* = 22,139)

|  |  | Uncertain |  |  | Unwilling |  |  |
| --- | --- | --- | --- | --- | --- | --- | --- |
|  |  | RRR | 95% CI |  | RRR | 95% CI |  |
| Gender (ref male) |  |  |  |  |  |  |  |
|  | Female | 1.06 | 0.74 | 1.52 | 0.96 | 0.61 | 1.50 |
| Age (ref 60+) |  |  |  |  |  |  |  |
|  | 45-59 | 1.60 | 1.00 | 2.54 | 1.57 | 0.91 | 2.72 |
|  | 30-44 | 4.31 | 2.63 | 7.05 | 2.56 | 1.31 | 5.00 |
|  | 18-29 | 5.30 | 2.87 | 9.77 | 5.74 | 2.82 | 11.68 |
| Ethnicity (ref white) |  |  |  |  |  |  |  |
|  | Ethnic minority groups | 1.07 | 0.58 | 1.95 | 1.50 | 0.74 | 3.05 |
| Education (ref undergraduate degree or higher) |  |  |  |  |  |  |  |
|  | A-levels or vocational | 1.51 | 0.95 | 2.39 | 2.09 | 1.15 | 3.79 |
|  | Up to GCSE | 1.76 | 1.09 | 2.84 | 2.50 | 1.31 | 4.79 |
| Income (ref >£90,000) |  |  |  |  |  |  |  |
|  | £60,000-89,999 | 0.50 | 0.21 | 1.22 | 0.35 | 0.11 | 1.07 |
|  | £30,000-59,999 | 2.43 | 1.06 | 5.59 | 0.58 | 0.22 | 1.54 |
|  | £16,000-29,999 | 2.25 | 0.88 | 5.78 | 1.38 | 0.49 | 3.87 |
|  | <£16,000 | 3.48 | 1.32 | 9.16 | 2.20 | 0.80 | 6.03 |
| Employment status (ref not employed) |  |  |  |  |  |  |  |
|  | Employed | 3.25 | 2.11 | 5.02 | 1.57 | 0.92 | 2.68 |
| Living arrangement (ref alone) |  |  |  |  |  |  |  |
|  | Living with others, not including children | 0.77 | 0.43 | 1.39 | 0.77 | 0.42 | 1.39 |
|  | Living with others, including children | 1.74 | 0.95 | 3.20 | 1.67 | 0.91 | 3.09 |
| Area of dwelling (ref urban) |  |  |  |  |  |  |  |
|  | Rural | 0.87 | 0.57 | 1.31 | 1.11 | 0.65 | 1.88 |
| Smoking status (ref former or non-smoker) |  |  |  |  |  |  |  |
|  | Current smoker | 1.61 | 0.96 | 2.72 | 1.08 | 0.57 | 2.05 |
| Key worker status (ref not a key worker) |  |  |  |  |  |  |  |
|  | Key worker | 0.77 | 0.49 | 1.21 | 1.26 | 0.70 | 2.24 |
| Long-term physical health condition (ref condition) |  |  |  |  |  |  |  |
|  | No long-term physical health condition | 1.67 | 1.17 | 2.40 | 1.52 | 1.00 | 2.30 |
| Long-term mental health condition (ref condition) |  |  |  |  |  |  |  |
|  | No long-term mental health condition | 1.19 | 0.80 | 1.79 | 1.43 | 0.82 | 2.50 |

Note. 'Very likely' to accept a COVID-19 booster vaccine was the reference group in the multinomial regression model. Ethnic minority groups refers to Black, Asian and minority ethnicity. GCSE refers to General Certificate of Secondary Education. Data were weighted to the proportions of gender, age, ethnicity, country, and education obtained from the Office for National Statistics.

For booster uncertainty, the same three factors continued to be predictors. Relative to adults aged 60 and above, adults aged 18-29 were over five times more likely to be uncertain (RRR = 5· 30; 95% CI: 2· 87 to 9· 77), adults aged 30-44 were over four times more likely (RRR = 4· 31; 95% CI: 2· 63 to 7· 05), and even adults aged 45-59 were 1· 6 times more likely to be uncertain (RRR = 1· 60; 95% CI: 1· 00 to 2· 54). Education up to GCSE was additionally associated with a 1· 7 times increased risk of being uncertain (RRR = 1· 76; 95% CI: 1· 09 to 2· 84) relative to having a degree, but the association for adults with A-levels or vocational qualifications was not present. As for unwillingness, healthy adults were also at increased risk of being unwilling (RRR = 1· 67; 95% CI: 1· 17 to 2· 40) to receive the booster compared to individuals with a long-term physical health condition. For uncertainty, two further socio-demographic factors emerged as predictors too. Adults who were employed (vs unemployed/retired/students/not working) were more than three times more likely to be uncertain (RRR = 3· 25; 95% CI: 2· 11 to 5· 02). There were mixed results for income, with some evidence that household income levels below £60,000 and below £16,000 per year were associated with increased risk of being uncertain (£30,000-59,999: RRR = 2· 43; 95% CI: 1· 06 to 5· 59; <£16,000: RRR = 3· 48; 95% CI: 1· 32 to 9· 16) relative to higher income households. But no further socio-demographic factors were associated with uncertainty, including ethnicity, living arrangement, smoking status, keyworker status and having a mental health condition were not associated with uncertainty or unwillingness.

### COVID-19 related predictors of booster uncertainty and unwillingness

The strongest predictor of booster uncertainty and unwillingness was original intentions to be vaccinated when the vaccines were first approved (Table 4). Adults who were initially uncertain about receiving a COVID-19 vaccine were five times more likely to say they were uncertain or unwilling to receive a booster (uncertain: RRR = 4· 92; 95% CI: 2· 98 to 8· 11; unwilling: RRR = 5· 29; 95% CI: 3· 07 to 9· 09) than those who were initially willing. Initial unwillingness associated with an over six times higher relative risk (RRR = 6· 40; 95% CI: 3· 94 to 10· 41) and eleven times higher relative risk (RRR = 11· 29; 95% CI: 6· 79 to 18· 78) of booster uncertainty and unwillingness, respectively.

**Table 4.** COVID-19 related predictors of uncertainty and unwillingness to receive a COVID-19 booster vaccine using a multivariable multinomial regression (weighted, *N* = 22,139)

|  | Uncertain |  |  | Unwilling |  |  |
| --- | --- | --- | --- | --- | --- | --- |
|  | RRR | 95% CI |  | RRR | 95% CI |  |
| Confidence in government to handle pandemic (ref high) |  |  |  |  |  |  |
| Low confidence in government to handle pandemic | 1.19 | 0.81 | 1.75 | 0.87 | 0.53 | 1.44 |
| Confidence in health system to handle pandemic (ref high) |  |  |  |  |  |  |
| Low confidence in health system to handle pandemic | 1.32 | 0.90 | 1.94 | 0.85 | 0.54 | 1.34 |
| Knowledge of COVID 19 (ref high) |  |  |  |  |  |  |
| Low knowledge of COVID-19 | 1.78 | 1.19 | 2.67 | 1.19 | 0.70 | 2.05 |
| Compliance across strict restriction periods (ref high) |  |  |  |  |  |  |
| Consistently low compliance with COVID-19 guidelines | 1.51 | 1.04 | 2.19 | 2.45 | 1.65 | 3.65 |
| Have had COVID-19 (ref have not had COVID-19) |  |  |  |  |  |  |
| Have had COVID-19, suspected by self or doctor | 1.38 | 0.89 | 2.15 | 1.24 | 0.76 | 2.03 |
| Have had COVID-19, confirmed by test | 1.11 | 0.66 | 1.88 | 1.06 | 0.59 | 1.91 |
| Stressed in the past week about catching/becoming ill from COVID-19 (stressed) |  |  |  |  |  |  |
| Not stressed | 1.80 | 1.19 | 2.72 | 1.91 | 1.19 | 3.07 |
| Initial COVID-19 vaccine intention (ref willing) |  |  |  |  |  |  |
| Initially uncertain about accepting a COVID-19 vaccine | 4.92 | 2.98 | 8.11 | 5.29 | 3.07 | 9.09 |
| Initially unwilling to accept a COVID-19 vaccine | 6.40 | 3.94 | 10.41 | 11.29 | 6.79 | 18.78 |

Note. 'Very likely' to accept a COVID-19 booster vaccine was the reference group in the multinomial regression model. Ethnic minority groups refers to Black, Asian and minority ethnicity. GCSE refers to General Certificate of Secondary Education. Data were weighted to the proportions of gender, age, ethnicity, country, and education obtained from the Office for National Statistics. Model includes all socio-demographic variables in Table 3.

The second highest predictor of unwillingness was consistently low compliance with COVID-19 guidelines during strict restriction periods (RRR = 2· 45; 95% CI: 1· 65 to 3· 65) which also predicted uncertainty (RRR = 1· 51; 95% CI: 1· 04 to 2· 19). Not having said that catching or falling seriously ill from COVID-19 was a source of stress in past week at follow-up also predicted unwillingness (RRR = 1· 91; 95% CI: 1· 19 to 3· 07) and uncertainty (RRR = 1· 80; 95% CI: 1· 19 to 2· 72) about boosters. Low self-reported knowledge of COVID-19 also predicted uncertainty but not unwillingness to get a booster (uncertainty: RRR = 1· 78; 95% CI: 1· 19 to 2· 67; unwillingness: RRR = 1· 19; 95% CI: 0· 70 to 2· 05). Confidence in government, confidence in the health system, and past experience of having COVID were not predictors of booster intentions.

### Sensitivity analyses

Sensitivity analyses conducted using participants’ last recorded (rather than first recorded) COVID-19 vaccine intention during the period 2 December 2020 to 31 March 2021 (Supplemental Table S5), from the model which included a variable indicating low vs high levels of compliance with guidelines across the entirety of the pandemic (rather than only during periods of strict restrictions) (Supplemental Table S6), and from the model which included only sample participants who had had exactly two doses of a COVID-19 vaccine at follow-up (Supplemental Table S7) resulted in substantively similar findings. However, amongst adults who had had exactly two doses of a COVID-19 vaccine at follow-up, age was no longer a predictor of booster uncertainty (likely because most older adults had already received boosters by then), whilst education up to A-levels or vocational and age 30-44 were no longer predictors of booster unwillingness (Supplemental Table S7).

### Role of the funding source

The funders had no role in the study design; in the collection, analysis, and interpretation of data; in the writing of the report; or in the decision to submit the paper for publication. All researchers listed as authors are independent from the funders and all final decisions about the research were taken by the investigators and were unrestricted. All authors had full access to all the data in the study and had final responsibility for the decision to submit for publication.

## Discussion

This is the first study, to our knowledge, to examine associations between socio-demographic and COVID-19 related factors, and previously measured initial intent to receive a COVID-19 vaccine with future uncertainty and unwillingness to receive a COVID-19 booster vaccine in fully vaccinated adults. The proportions in our sample who were uncertain about receiving (4%) and unwilling (4%) to receive a COVID-19 booster vaccine are similar to those in the OPN (4% and 5%, respectively).^18^ Our findings broadly align with the factors already identified as predicting initial vaccine intentions when the vaccine was first approved (e.g. age, income, employment, and education),^11,15,24,25^ showing that some of the same groups should remain the focus of public health campaigns to increase booster vaccine uptake. Underlining this finding, our results also highlight that previous uncertainty and unwillingness to be vaccinated remains the strongest predictor of intentions to receive a booster vaccine, even amongst adults who did get fully vaccinated despite initial concerns. Two other concerning patterns also emerged from our results. First, adults from lower socio-economic backgrounds were most likely to be uncertain or unwilling to accept a booster vaccine as well as most likely to be seriously affected by the virus. Unequal uptake of booster vaccinations may therefore increase social inequalities in experiences of COVID-19. Second, some of those most likely to spread COVID-19 (i.e., those with poor compliance with guidelines) are most likely to be uncertain and unwilling.

Previous vaccine attitudes and intentions are logical predictors of attitudes to booster vaccines, and previous research has highlighted a range of reasons for initial hesitancy, including high levels of mistrust in vaccines and concerns about unforeseen future side effects,^11^ as well as a belief that immunity through infection is safest and that the COVID-19 vaccines were insufficiently tested.^15^ However, the adults in our study had already received at least two vaccines, so their initial uncertainty or unwillingness had previously been overcome. Furthermore, not all of the fully vaccinated adults in our sample who were uncertain or unwilling to receive a booster vaccine were initially hesitant about receiving a first dose (Table 2); 5% of the initially willing were now either uncertain (3%) or unwilling (2%) to accept a booster vaccine. So, there is a need to understand the reasons for uncertainty and unwillingness to accept a booster vaccine in more detail. One potential explanation has to do with misinformation about the length of protection provided by COVID-19 vaccines. In recent data from fully vaccinated adults in the OPN study, the most commonly cited reason for unwillingness to receive a booster vaccine (59%) was a belief that the first and second vaccines were enough to keep them safe, and nearly half (49%) thought that a booster would not offer them extra protection.^18^ Indeed, it is notable that healthy adults were 1.5 times more likely to be uncertain or unwilling to receive a booster vaccine, suggesting that the booster vaccine is held as less important than individuals with pre-existing health conditions. This indicates an urgent need for public health messaging to correct widespread misinformation on the length of protection offered from COVID-19 vaccines. A second potential explanation relates to recent publicised concerns that giving third doses to people in high income countries whilst many lower income countries do not have access to a first or second dose will increase inequities.^26^ Indeed, recent findings from the OPN indicate that around one in four (22%) fully vaccinated adults who are unwilling to accept a booster vaccine said they were unwilling to do so because they thought the vaccine should be offered to someone else.^18^ Further, a study conducted by Imperial College London asked respondents specifically if they would be willing to donate their COVID-19 booster vaccine to people in low income countries if a test showed they did not need a booster dose, and the majority of respondents in the UK (78%) and the US (69%) said they would do so.^27^ Therefore, altruism may be a driver for booster hesitancy or unwillingness for some adults in our sample. Further research is needed to understand these new attitudes further in order that specific strategies to encourage booster uptake can be designed and implemented.

It is also concerning that some we found a socio-economic gradient in intentions to take up the booster vaccine, as some of the same socio-demographic groups most at risk of uncertainty and unwillingness to receive the COVID-19 booster vaccine are also at increased risk for severe illness and death from COVID-19, including those with low incomes and who have low levels of education.^11,13,14,28^ These groups have already experienced a disproportionally higher number of infections and deaths, relative to other groups and had lower uptake of initial vaccines.^29–32^ Consequently, our finding of reduced intentions to take up the booster in this group, even amongst individuals who have already received two vaccinations, is concerning as it may exacerbate existing inequalities in exposure to and experience of the virus. However, other socio-demographic groups did not show any differences in booster vaccine intentions (key worker status, smoking, ethnicity etc.) This provides direction as to which groups may require more specific public health strategies to encourage booster vaccinations, and such work may need to take account of specific logistical or financial barriers individuals may face in getting their booster vaccine.

Worryingly, some of those who are most likely to spread the virus are also the least likely to say they will accept a booster vaccine. First, the most important socio-demographic determinant of both uncertainty and unwillingness to receive a COVID-19 booster vaccine in our study was age, which has been found in prior work.^13,18,28^ Young adults were over five times as likely to be uncertain about accepting a booster vaccine compared to older adults, and nearly six times as likely to be unwilling. Young adults may be more hesitant about accepting a booster vaccine because they feel they are less likely to be severely affected by the disease. Other work has shown that young adults are less stressed by the virus^33^ and less compliant with government guidelines,^34^ which were also factors identified as predictors of original COVID-19 vaccine unwillingness and hesitancy.^11,28^ Young people may also be less likely to be vaccinated because of concerns about side effects.^11^ However, in the OPN study, young adults (and adolescents) who were initially uncertain about accepting a COVID-19 vaccine were the most likely to have later received at least one dose.^20^ Therefore, the younger adults age group may be most amenable to public health programmes to provide more information and clarity around the benefits of booster vaccination. We also found an association between uncertainty and unwillingness to receive a COVID-19 booster vaccine and poor compliance with government guidelines during lockdowns, which is congruent with other research.^11,28^ This is concerning, as individuals who are non-compliant with behavioural interventions such as mask wearing and isolation when infected or symptomatic, or after contact with someone who has been infected, are more likely to spread the virus.^35^ Hospitalisations may subsequently increase, further driving the need for more of the behavioural interventions these individuals fail to comply with.

Interestingly, though, having been previously infected with COVID-19 was not a predictor of either unwillingness or uncertainty about accepting a booster vaccine, which was also found in a large multi-national survey in relation to COVID-19 vaccine intent.^24^ However, in prior work using data from the current study, respondents who had previously been infected with COVID-19 were more likely to be uncertain about accepting, but not unwilling to accept a COVID-19 vaccine,^11^ and in a national survey conducted in early 2021 in the UK, respondents who had ‘definitely’ or ‘probably’ had COVID-19 were less likely to say they would accept a vaccine.^36^ Prior COVID-19 infection was also associated with reduced likelihood of vaccine uptake in the Coronavirus (COVID-19) Infection Survey in October 2021.^10^ This suggests that individuals who have not yet received a first dose may be making a trade in terms of strategies to avoid infection, or are unaware of how long COVID-19 infection confers immunity. Future research should examine how factors such as severity of infection and timing of infection relative to receiving a COVID-19 vaccine relate to booster vaccine intentions in fully vaccinated adults.

Finally, it is notable that we found that some people willing to accept a booster had initially been uncertain or unwilling to have a vaccination when the vaccines were first approved in 2020. As our sample was restricted to people who had received two vaccinations, it only covers individuals who had overcome initial uncertainty or unwillingness to be vaccinated. Nonetheless, 73% of those who were initially uncertain and 65% of those who were initially unwilling to be vaccinated but who had still received two vaccinations were willing to accept a booster. This highlights that previous vaccination attitudes can be changed not just for single vaccinations but for multiple courses of vaccination. It supports previous research suggesting that people who were initially uncertain rather than unwilling may be easier to convince to vaccinate than the latter, and therefore a promising group to engage in health communication programmes.^37^ For example, the Vaccine Opinions Study in the UK, adults who were previously unsure about the COVID-19 vaccine were more likely to later have accepted at least one dose it than those who were initially unwilling (60% vs 40%).^20^ However, our data still suggest there could be a value to trying to engage people who are supposedly unwilling. We recommend that more research is undertaken to ascertain why these groups changed their minds about being vaccinated as such findings could provide valuable insights for public health messaging.

### Strengths and limitations

Strengths of the current study are that it is a large sample well-stratified on major demographic characteristics and analyses were weighted to increase representativeness of the general UK population. Analyses were also longitudinal and included rich information on a range of socio-demographic and COVID-19 related factors. We focused on initial COVID-19 vaccine attitudes in the four months following the day their approval was announced in the UK in 2019 as well as gathering new data on booster vaccine attitudes to allow comparisons of attitudes over the past twelve months. However, there were some limitations. First, our sample is not nationally representative of the general UK population. We used a heterogeneous and large sample in our analyses and weighted our responses to population proportions, but it is possible that more extreme views towards booster vaccines may therefore not be represented. Second, our recruitment strategy could have resulted in clustering (i.e., individuals from the same households participating), and attitudes may be more similar within households. However, this would be considered a minimal risk given the large sample and we used robust standard errors. Third, due to small sub-group sample sizes we were unable to examine specific effects of different ethnicities on booster vaccine attitudes and we treated this socio-demographic characteristic as binary. This could have led to oversimplification of the diverse opinions and experiences held by different ethnicity subgroups.

## Conclusions

Findings suggest initial uncertainty and unwillingness about receiving a COVID-19 booster vaccine will be a barrier to maintaining the progress that has been made thus far in controlling the COVID-19 pandemic through vaccines. There is therefore an urgent need for public health messaging emphasising the safety and importance of COVID-19 booster vaccines even for fully vaccinated adults, in particular focusing on individuals from lower socio-economic backgrounds and individuals at greatest risk of spreading the virus. Finally, in light of evidence showing that existing COVID-19 vaccines offer protection for limited time periods and therefore will likely need to be updated on a regular period moving forwards, there is a need for more research into ongoing attitudes to COVID-19 vaccinations and boosters.

## Data Availability

The UCL COVID-19 Social Study documentation and codebook are available for download at https://www.covidsocialstudy.org/. Statistical code is available upon request from Elise Paul.

## Contributors

DF conceptualised and designed the study. DF also acquired funding, led the investigation, provided oversight on the methodology, administered the project, provided software and other resources, and supervised the project. Data were curated, validated, and formally analysed by EP. EP created visualisations, wrote the original manuscript draft with input from all authors. Both authors reviewed and edited the manuscript. Both authors approved the final version of the manuscript and had full access to and verified the data.

## Acknowledgements

The researchers are grateful for the support of a number of organisations with their recruitment efforts including: the UKRI Mental Health Networks, Find Out Now, UCL BioResource, SEO Works, FieldworkHub, and Optimal Workshop.

## Data availability statement

The UCL COVID-19 Social Study documentation and codebook are available for download at https://osf.io/jm8ra/. Statistical code is available upon request from Elise Paul.

## Ethics approval and consent to participate

Ethical approval for the COVID-19 Social Study was granted by the UCL Ethics Committee. All participants provided fully informed consent and the study is GDPR compliant.

## Supplementary Material

**Table S1.** Comparison of excluded and included participants, unweighted.

|  |  | Excluded |  | Included |  |
| --- | --- | --- | --- | --- | --- |
|  |  | Mean | SD | Mean | SD |
| Gender | Male | 0.23 | 0.42 | 0.26 | 0.44 |
|  | Female | 0.77 | 0.42 | 0.74 | 0.44 |
| Age | 60+ | 0.36 | 0.48 | 0.43 | 0.50 |
|  | 45-59 | 0.35 | 0.48 | 0.34 | 0.47 |
|  | 30-44 | 0.22 | 0.42 | 0.19 | 0.39 |
|  | 18-29 | 0.07 | 0.25 | 0.04 | 0.19 |
| Ethnicity | White | 0.95 | 0.21 | 0.97 | 0.18 |
|  | Ethnic minority groups | 0.05 | 0.21 | 0.03 | 0.18 |
| Education | Undergraduate degree or higher | 0.64 | 0.48 | 0.71 | 0.46 |
|  | A-levels or vocational | 0.19 | 0.39 | 0.16 | 0.37 |
|  | Up to GCSE | 0.17 | 0.38 | 0.13 | 0.34 |
| Income | >£90,000 | 0.09 | 0.29 | 0.10 | 0.30 |
|  | £60,000-89,999 | 0.15 | 0.35 | 0.15 | 0.35 |
|  | £30,000-59,999 | 0.34 | 0.47 | 0.35 | 0.48 |
|  | £16,000-29,999 | 0.25 | 0.43 | 0.26 | 0.44 |
|  | <£16,000 | 0.17 | 0.38 | 0.14 | 0.35 |
| Employment status | Not employed | 0.41 | 0.49 | 0.42 | 0.49 |
|  | Employed | 0.59 | 0.49 | 0.58 | 0.49 |
| Living arrangement | Alone | 0.18 | 0.38 | 0.23 | 0.42 |
|  | With others, not with children | 0.56 | 0.50 | 0.57 | 0.49 |
|  | With, including children | 0.26 | 0.44 | 0.20 | 0.40 |
| Area of dwelling | Urban | 0.74 | 0.44 | 0.75 | 0.43 |
|  | Rural | 0.26 | 0.44 | 0.25 | 0.43 |
| Smoking status | Former or non-smoker | 0.90 | 0.30 | 0.93 | 0.25 |
|  | Current smoker | 0.10 | 0.30 | 0.07 | 0.25 |
| Key worker status | Not a key worker | 0.76 | 0.43 | 0.80 | 0.40 |
|  | Key worker | 0.24 | 0.43 | 0.20 | 0.40 |
| Long-term physical health condition | No long-term physical health condition | 0.57 | 0.50 | 0.57 | 0.50 |
|  | Long-term physical health condition | 0.43 | 0.50 | 0.43 | 0.50 |
| Long-term mental health condition | No long-term mental health condition | 0.81 | 0.39 | 0.85 | 0.36 |
|  | Long-term mental health condition | 0.19 | 0.39 | 0.15 | 0.36 |
| Confidence in government to handle pandemic |  |  |  |  |  |
|  | Low confidence in government to handle pandemic | 0.31 | 0.46 | 0.33 | 0.47 |
|  | Low confidence in government to handle pandemic | 0.69 | 0.46 | 0.67 | 0.47 |
| Confidence in health system to handle pandemic |  |  |  |  |  |
|  | High confidence in health system to handle pandemic | 0.52 | 0.50 | 0.62 | 0.49 |
|  | Low confidence in health system to handle pandemic | 0.48 | 0.50 | 0.38 | 0.49 |
| Knowledge of COVID 19 |  |  |  |  |  |
|  | High knowledge of COVID-19 | 0.86 | 0.35 | 0.87 | 0.34 |
|  | Low knowledge of COVID-19 | 0.14 | 0.35 | 0.13 | 0.34 |
| Compliance across strict restriction periods |  |  |  |  |  |
|  | Consistently high compliance with COVID-19 guidelines | 0.75 | 0.44 | 0.72 | 0.45 |
|  | Consistently low compliance with COVID-19 guidelines | 0.25 | 0.44 | 0.28 | 0.45 |
| Have had COVID-19 |  |  |  |  |  |
|  | Have not had COVID-19 | 0.74 | 0.44 | 0.79 | 0.41 |
|  | Have had COVID-19, suspected by self or doctor | 0.12 | 0.32 | 0.10 | 0.30 |
|  | Have had COVID-19, confirmed by test | 0.14 | 0.34 | 0.11 | 0.31 |
| Stressed in the past week about catching/becoming ill from COVID-19 |  |  |  |  |  |
|  | Stressed | 0.32 | 0.47 | 0.35 | 0.48 |
|  | Not stressed | 0.68 | 0.47 | 0.65 | 0.48 |
| Initial COVID-19 vaccine intention |  |  |  |  |  |
|  | Initially willing to accept or had accepted a COVID-19 vaccine | 0.80 | 0.40 | 0.92 | 0.27 |
|  | Initially uncertain about accepting a COVID-19 vaccine | 0.08 | 0.26 | 0.05 | 0.22 |
|  | Initially unwilling to accept a COVID-19 vaccine | 0.12 | 0.33 | 0.03 | 0.16 |
| COVID-19 booster vaccine intentions |  |  |  |  |  |
|  | Willing/accepted a COVID-19 booster vaccine | 0.85 | 0.36 | 0.95 | 0.21 |
|  | Uncertain about a COVID-19 booster vaccine | 0.04 | 0.19 | 0.02 | 0.16 |
|  | Unwilling to accept COVID-19 booster vaccine | 0.11 | 0.32 | 0.02 | 0.15 |

Note. 'Very likely' to accept a COVID-19 booster vaccine was the reference group in the multinomial regression model. Ethnic minority groups refers to Black, Asian and minority ethnicity. GCSE refers to General Certificate of Secondary Education.

**Table S2.** Wording of study developed items.

| Variable | Question wording | Response options |
| --- | --- | --- |
| Ethnicity | What is your ethnicity? | 1- Asian/Asian British - Indian, Pakistani, Bangladeshi, other |
|  |  | 2- Black/Black British - Caribbean, African, other |
|  |  | 3- Mixed race - White and Black/Black British |
|  |  | 4- Mixed race – other |
|  |  | 5- White - British, Irish, other |
|  |  | 6- Chinese/Chinese British |
|  |  | 7- Middle Eastern/Middle Eastern British - Arab, Turkish, other |
|  |  | 8- Other ethnic group |
|  |  | 9- Prefer not to say |
| Intention to receive a COVID-19 booster vaccine | We are interested in your thoughts on the BOOSTER VACCINATION. | 1- Very unlikely |
|  | How likely do you think you are to get a COVID-19 booster vaccine if/when you are offered one? | 6- very likely |
|  |  | I have already had one/accepted one |
| Have ever had COVID-19 | Overall, do you believe you have ever had Covid-19? | Yes, confirmed by a positive Covid-19 test at the time I felt ill |
|  |  | Yes, confirmed by a positive antibody test since |
|  |  | Yes, suspected by a doctor but not tested |
|  |  | Yes, my own suspicions |
|  |  | No, not that I know of |
| Confidence in government to handle pandemic | To what extent do you have confidence in the UK Government's ability to handle the Coronavirus situation as it continues to develop? | 1- none at all to |
|  |  | 7- lots |
| Confidence in health system to handle pandemic | How much confidence do you have that the UK health service can cope during Covid-19? If you live in a devolved nation, we ask you to focus on the health service within your country (e.g., NHS Health Scotland / NHS Wales / HSCNI)? | 1- none at all to |
|  |  | 7- lots |
| Compliance with COVID-19 guidelines | Are you following the recommendations from authorities to prevent spread of COVID-19? | 1- none at all to |
|  |  | 7- very much so |
| Knowledge of COVID-19 | How would you rate your knowledge of COVID-19? | 1- very poor knowledge to |
|  |  | 7- very good knowledge |

**Table S3.** Dates of strict restrictions in England.

| **Restriction period** | **Dates** |
| --- | --- |
| First | 23 March to 9 May 2020 |
| Second | 2 November to 3 December 2020 |
| Third | 5 January to 12 April 2021 |

**Table S4.**
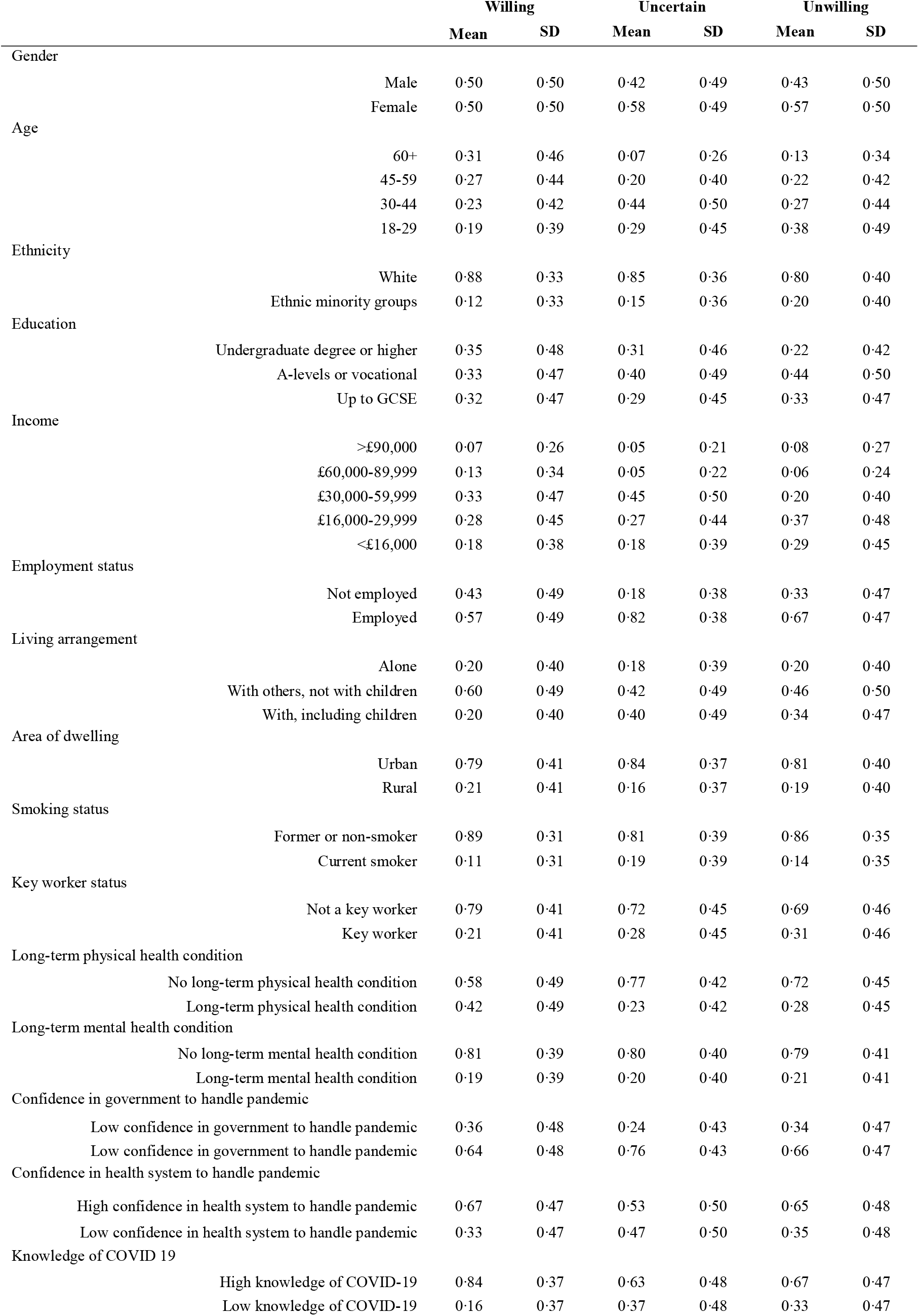
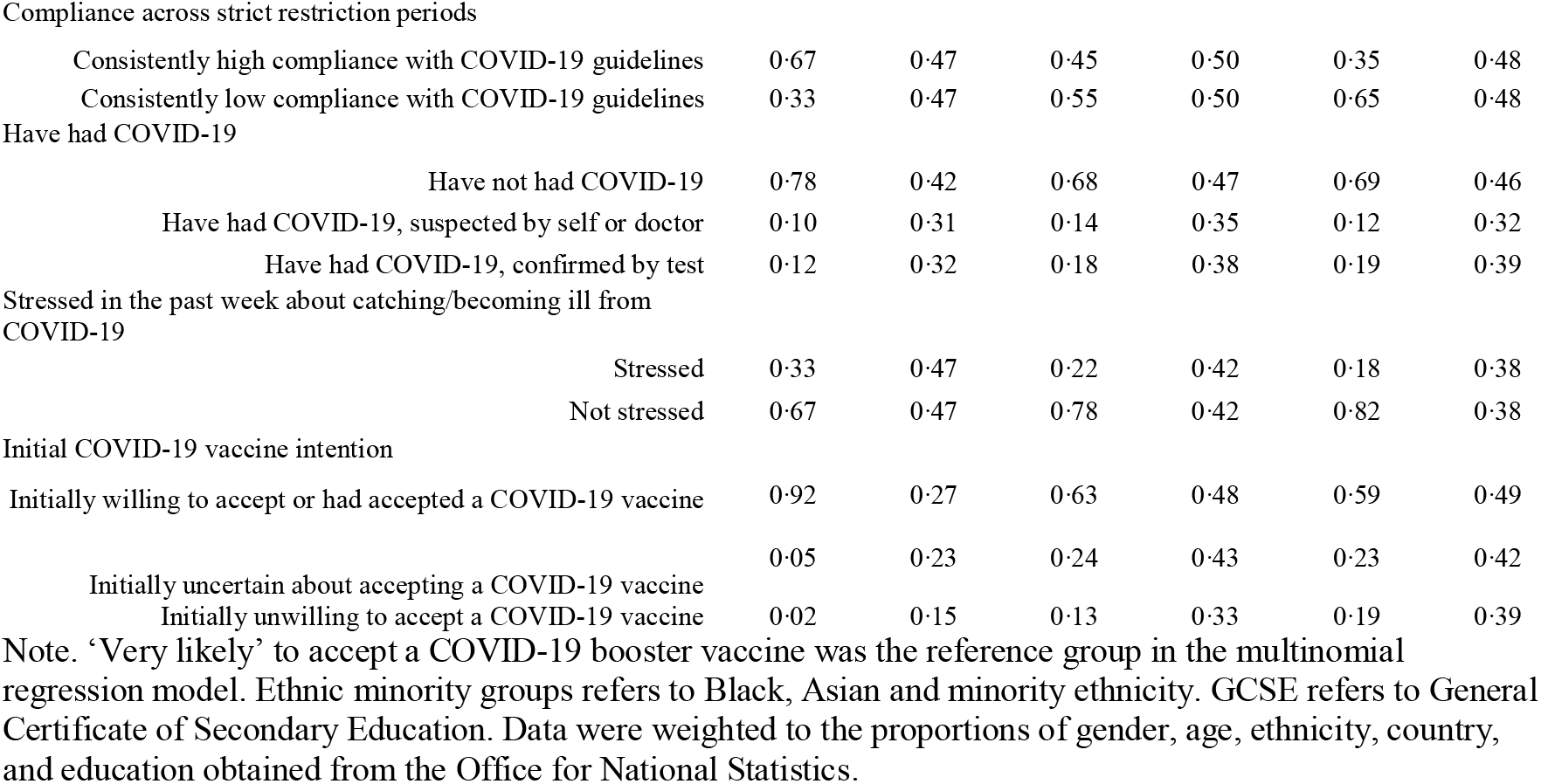
Comparison of sample participants by COVID-19 booster vaccine intention (weighted, *N* = 22,139)

|  |  | Willing |  | Uncertain |  | Unwilling |  |
| --- | --- | --- | --- | --- | --- | --- | --- |
|  |  | Mean | SD | Mean | SD | Mean | SD |
| Gender |  |  |  |  |  |  |  |
|  | Male | 0.50 | 0.50 | 0.42 | 0.49 | 0.43 | 0.50 |
|  | Female | 0.50 | 0.50 | 0.58 | 0.49 | 0.57 | 0.50 |
| Age |  |  |  |  |  |  |  |
|  | 60+ | 0.31 | 0.46 | 0.07 | 0.26 | 0.13 | 0.34 |
|  | 45-59 | 0.27 | 0.44 | 0.20 | 0.40 | 0.22 | 0.42 |
|  | 30-44 | 0.23 | 0.42 | 0.44 | 0.50 | 0.27 | 0.44 |
|  | 18-29 | 0.19 | 0.39 | 0.29 | 0.45 | 0.38 | 0.49 |
| Ethnicity |  |  |  |  |  |  |  |
|  | White | 0.88 | 0.33 | 0.85 | 0.36 | 0.80 | 0.40 |
|  | Ethnic minority groups | 0.12 | 0.33 | 0.15 | 0.36 | 0.20 | 0.40 |
| Education |  |  |  |  |  |  |  |
|  | Undergraduate degree or higher | 0.35 | 0.48 | 0.31 | 0.46 | 0.22 | 0.42 |
|  | A-levels or vocational | 0.33 | 0.47 | 0.40 | 0.49 | 0.44 | 0.50 |
|  | Up to GCSE | 0.32 | 0.47 | 0.29 | 0.45 | 0.33 | 0.47 |
| Income |  |  |  |  |  |  |  |
|  | >£90,000 | 0.07 | 0.26 | 0.05 | 0.21 | 0.08 | 0.27 |
|  | £60,000-89,999 | 0.13 | 0.34 | 0.05 | 0.22 | 0.06 | 0.24 |
|  | £30,000-59,999 | 0.33 | 0.47 | 0.45 | 0.50 | 0.20 | 0.40 |
|  | £16,000-29,999 | 0.28 | 0.45 | 0.27 | 0.44 | 0.37 | 0.48 |
|  | <£16,000 | 0.18 | 0.38 | 0.18 | 0.39 | 0.29 | 0.45 |
| Employment status |  |  |  |  |  |  |  |
|  | Not employed | 0.43 | 0.49 | 0.18 | 0.38 | 0.33 | 0.47 |
|  | Employed | 0.57 | 0.49 | 0.82 | 0.38 | 0.67 | 0.47 |
| Living arrangement |  |  |  |  |  |  |  |
|  | Alone | 0.20 | 0.40 | 0.18 | 0.39 | 0.20 | 0.40 |
|  | With others, not with children | 0.60 | 0.49 | 0.42 | 0.49 | 0.46 | 0.50 |
|  | With, including children | 0.20 | 0.40 | 0.40 | 0.49 | 0.34 | 0.47 |
| Area of dwelling |  |  |  |  |  |  |  |
|  | Urban | 0.79 | 0.41 | 0.84 | 0.37 | 0.81 | 0.40 |
|  | Rural | 0.21 | 0.41 | 0.16 | 0.37 | 0.19 | 0.40 |
| Smoking status |  |  |  |  |  |  |  |
|  | Former or non-smoker | 0.89 | 0.31 | 0.81 | 0.39 | 0.86 | 0.35 |
|  | Current smoker | 0.11 | 0.31 | 0.19 | 0.39 | 0.14 | 0.35 |
| Key worker status |  |  |  |  |  |  |  |
|  | Not a key worker | 0.79 | 0.41 | 0.72 | 0.45 | 0.69 | 0.46 |
|  | Key worker | 0.21 | 0.41 | 0.28 | 0.45 | 0.31 | 0.46 |
| Long-term physical health condition |  |  |  |  |  |  |  |
|  | No long-term physical health condition | 0.58 | 0.49 | 0.77 | 0.42 | 0.72 | 0.45 |
|  | Long-term physical health condition | 0.42 | 0.49 | 0.23 | 0.42 | 0.28 | 0.45 |
| Long-term mental health condition |  |  |  |  |  |  |  |
|  | No long-term mental health condition | 0.81 | 0.39 | 0.80 | 0.40 | 0.79 | 0.41 |
|  | Long-term mental health condition | 0.19 | 0.39 | 0.20 | 0.40 | 0.21 | 0.41 |
| Confidence in government to handle pandemic |  |  |  |  |  |  |  |
|  | Low confidence in government to handle pandemic | 0.36 | 0.48 | 0.24 | 0.43 | 0.34 | 0.47 |
|  | High confidence in government to handle pandemic | 0.64 | 0.48 | 0.76 | 0.43 | 0.66 | 0.47 |
| Confidence in health system to handle pandemic |  |  |  |  |  |  |  |
|  | High confidence in health system to handle pandemic | 0.67 | 0.47 | 0.53 | 0.50 | 0.65 | 0.48 |
|  | Low confidence in health system to handle pandemic | 0.33 | 0.47 | 0.47 | 0.50 | 0.35 | 0.48 |
| Knowledge of COVID 19 |  |  |  |  |  |  |  |
|  | High knowledge of COVID-19 | 0.84 | 0.37 | 0.63 | 0.48 | 0.67 | 0.47 |
|  | Low knowledge of COVID-19 | 0.16 | 0.37 | 0.37 | 0.48 | 0.33 | 0.47 |

|  |  |  |  |  |  |  |
| --- | --- | --- | --- | --- | --- | --- |
| Compliance across strict restriction periods |  |  |  |  |  |  |
| Consistently high compliance with COVID-19 guidelines | 0.67 | 0.47 | 0.45 | 0.50 | 0.35 | 0.48 |
| Consistently low compliance with COVID-19 guidelines | 0.33 | 0.47 | 0.55 | 0.50 | 0.65 | 0.48 |
| Have had COVID-19 |  |  |  |  |  |  |
| Have not had COVID-19 | 0.78 | 0.42 | 0.68 | 0.47 | 0.69 | 0.46 |
| Have had COVID-19, suspected by self or doctor | 0.10 | 0.31 | 0.14 | 0.35 | 0.12 | 0.32 |
| Have had COVID-19, confirmed by test | 0.12 | 0.32 | 0.18 | 0.38 | 0.19 | 0.39 |
| Stressed in the past week about catching/becoming ill from COVID-19 |  |  |  |  |  |  |
| Stressed | 0.33 | 0.47 | 0.22 | 0.42 | 0.18 | 0.38 |
| Not stressed | 0.67 | 0.47 | 0.78 | 0.42 | 0.82 | 0.38 |
| Initial COVID-19 vaccine intention |  |  |  |  |  |  |
| Initially willing to accept or had accepted a COVID-19 vaccine | 0.92 | 0.27 | 0.63 | 0.48 | 0.59 | 0.49 |
| Initially uncertain about accepting a COVID-19 vaccine | 0.05 | 0.23 | 0.24 | 0.43 | 0.23 | 0.42 |
| Initially unwilling to accept a COVID-19 vaccine | 0.02 | 0.15 | 0.13 | 0.33 | 0.19 | 0.39 |

Note. 'Very likely' to accept a COVID-19 booster vaccine was the reference group in the multinomial regression model. Ethnic minority groups refers to Black, Asian and minority ethnicity. GCSE refers to General Certificate of Secondary Education. Data were weighted to the proportions of gender, age, ethnicity, country, and education obtained from the Office for National Statistics.

**Table S5.** Sensitivity analysis: Socio-demographic, COVID-19 related, and initial COVID-19 vaccine intent predictors of uncertainty and unwillingness to receive a COVID-19 booster vaccine using a multivariable multinomial regression and last recorded initial COVID-19 vaccine intent (weighted, *N* = 22,319)

|  |  | Uncertain |  |  | Unwilling |  |  |
| --- | --- | --- | --- | --- | --- | --- | --- |
|  |  | RRR | 95% CI |  | RRR | 95% CI |  |
| Gender (ref male) | Female | 1.12 | 0.77 | 1.62 | 1.06 | 0.68 | 1.65 |
| Age (ref 60+) | 45-59 | 1.43 | 0.90 | 2.25 | 1.33 | 0.77 | 2.28 |
|  | 30-44 | 3.09 | 1.82 | 5.25 | 1.61 | 0.81 | 3.21 |
|  | 18-29 | 3.20 | 1.76 | 5.83 | 3.24 | 1.46 | 7.23 |
| Ethnicity (ref white) | Ethnic minority groups | 0.80 | 0.42 | 1.50 | 1.13 | 0.53 | 2.42 |
| Education (ref undergraduate degree or higher) | A-levels or vocational | 1.39 | 0.87 | 2.20 | 1.69 | 0.98 | 2.92 |
|  | Up to GCSE | 1.57 | 1.00 | 2.45 | 2.03 | 1.14 | 3.59 |
| Income (ref >£90,000) | £60,000-89,999 | 0.49 | 0.20 | 1.23 | 0.34 | 0.12 | 0.99 |
|  | £30,000-59,999 | 2.54 | 1.10 | 5.87 | 0.69 | 0.29 | 1.66 |
|  | £16,000-29,999 | 2.27 | 0.89 | 5.79 | 1.41 | 0.53 | 3.78 |
|  | <£16,000 | 3.40 | 1.25 | 9.19 | 2.37 | 0.89 | 6.30 |
| Employment status (ref not employed) | Employed | 3.29 | 2.10 | 5.14 | 1.57 | 0.91 | 2.71 |
| Living arrangement (ref alone) | Living with others, not including children | 0.83 | 0.47 | 1.45 | 0.83 | 0.45 | 1.53 |
|  | Living with others, including children | 1.76 | 0.95 | 3.25 | 1.66 | 0.87 | 3.16 |
| Area of dwelling (ref urban) | Rural | 0.86 | 0.56 | 1.34 | 1.01 | 0.55 | 1.87 |
| Smoking status (ref former or non-smoker) | Current smoker | 1.65 | 1.02 | 2.68 | 1.22 | 0.63 | 2.35 |
| Key worker status (ref not a key worker) | Key worker | 0.74 | 0.49 | 1.13 | 1.20 | 0.67 | 2.15 |
| Long-term physical health condition (ref condition) | No long-term physical health condition | 1.41 | 0.98 | 2.05 | 1.09 | 0.71 | 1.68 |
| Long-term mental health condition (ref condition) | No long-term mental health condition | 1.34 | 0.87 | 2.06 | 1.52 | 0.83 | 2.79 |
| Confidence in government to handle pandemic (ref high) | Low confidence in government to handle pandemic | 1.19 | 0.81 | 1.76 | 0.85 | 0.51 | 1.42 |
| Confidence in health system to handle pandemic (ref high) | Low confidence in health system to handle pandemic | 1.32 | 0.91 | 1.93 | 0.83 | 0.53 | 1.30 |
| Knowledge of COVID 19 (ref high) | Low knowledge of COVID-19 | 1.81 | 1.22 | 2.68 | 1.11 | 0.63 | 1.95 |
| Compliance across strict restriction periods (ref high) | Consistently low compliance with COVID-19 guidelines | 1.58 | 1.09 | 2.31 | 2.82 | 1.87 | 4.25 |
| Have had COVID-19 (ref have not had COVID-19) | Have had COVID-19, suspected by self or doctor | 1.37 | 0.87 | 2.14 | 1.30 | 0.80 | 2.14 |
|  | Have had COVID-19, confirmed by test | 1.16 | 0.69 | 1.93 | 1.12 | 0.63 | 1.99 |
| Stressed in the past week about catching/becoming ill from COVID-19 (ref stressed) | Not stressed | 1.79 | 1.17 | 2.75 | 1.85 | 1.15 | 2.95 |
| Initial COVID-19 vaccine intention (ref willing) | Initially uncertain about accepting a COVID-19 vaccine | 4.41 | 2.08 | 9.35 | 10.86 | 5.18 | 22.78 |
|  | Initially unwilling to accept a COVID-19 vaccine | 6.47 | 2.46 | 17.02 | 10.67 | 3.98 | 28.59 |

Note. 'Very likely' to accept a COVID-19 booster vaccine was the reference group in the multinomial regression model. Ethnic minority groups refers to Black, Asian and minority ethnicity. GCSE refers to General Certificate of Secondary Education. Data were weighted to the proportions of gender, age, ethnicity, country, and education obtained from the Office for National Statistics.

**Table S6.** Sensitivity analysis: Socio-demographic, COVID-19 related, and initial COVID-19 vaccine intent predictors of uncertainty and unwillingness to receive a COVID-19 booster vaccine using a multivariable multinomial regression using average compliance with government guidelines across the entirety of the pandemic (weighted, *N* = 22,139)

|  |  | RRR | Uncertain<br>95% CI |  | RRR | Unwilling<br>95% CI |  |
| --- | --- | --- | --- | --- | --- | --- | --- |
| Gender (ref male) | Female | 1.05 | 0.72 | 1.53 | 0.98 | 0.64 | 1.50 |
| Age (ref 60+) | 45-59 | 1.45 | 0.91 | 2.31 | 1.46 | 0.83 | 2.56 |
|  | 30-44 | 3.33 | 1.97 | 5.64 | 2.05 | 1.01 | 4.16 |
|  | 18-29 | 3.91 | 2.14 | 7.12 | 4.73 | 2.12 | 10.52 |
| Ethnicity (ref white) | Ethnic minority groups | 0.77 | 0.40 | 1.48 | 1.18 | 0.59 | 2.35 |
| Education (ref undergraduate degree or higher) | A-levels or vocational | 1.33 | 0.84 | 2.13 | 1.78 | 1.04 | 3.06 |
|  | Up to GCSE | 1.47 | 0.93 | 2.30 | 2.12 | 1.20 | 3.77 |
| Income (ref >£90,000) | £60,000-89,999 | 0.50 | 0.20 | 1.26 | 0.34 | 0.11 | 1.06 |
|  | £30,000-59,999 | 2.52 | 1.08 | 5.85 | 0.70 | 0.28 | 1.74 |
|  | £16,000-29,999 | 2.22 | 0.87 | 5.70 | 1.39 | 0.52 | 3.71 |
|  | <£16,000 | 3.07 | 1.13 | 8.29 | 2.09 | 0.78 | 5.56 |
| Employment status (ref not employed) | Employed | 3.15 | 2.00 | 4.94 | 1.43 | 0.84 | 2.46 |
| Living arrangement (ref alone) | Living with others, not including children | 0.73 | 0.42 | 1.28 | 0.72 | 0.39 | 1.33 |
|  | Living with others, including children | 1.67 | 0.89 | 3.12 | 1.61 | 0.85 | 3.08 |
| Area of dwelling (ref urban) | Rural | 0.87 | 0.55 | 1.36 | 1.01 | 0.56 | 1.81 |
| Smoking status (ref former or non-smoker) | Current smoker | 1.79 | 1.11 | 2.91 | 1.27 | 0.66 | 2.44 |
| Key worker status (ref not a key worker) | Key worker | 0.74 | 0.48 | 1.14 | 1.21 | 0.67 | 2.18 |
| Long-term physical health condition (ref condition) | No long-term physical health condition | 1.40 | 0.96 | 2.04 | 1.18 | 0.77 | 1.79 |
| Long-term mental health condition (ref condition) | No long-term mental health condition | 1.37 | 0.88 | 2.13 | 1.54 | 0.86 | 2.75 |
| Confidence in government to handle pandemic (ref high) | Low confidence in government to handle pandemic | 1.19 | 0.81 | 1.75 | 0.87 | 0.53 | 1.44 |
| Confidence in health system to handle pandemic (ref high) | Low confidence in health system to handle pandemic | 1.32 | 0.90 | 1.94 | 0.85 | 0.54 | 1.34 |
| Knowledge of COVID 19 (ref high) | Low knowledge of COVID-19 | 1.78 | 1.19 | 2.67 | 1.19 | 0.70 | 2.05 |
| Compliance across strict restriction periods (ref high) | Consistently low compliance with COVID-19 guidelines | 1.51 | 1.04 | 2.19 | 2.45 | 1.65 | 3.65 |
| Have had COVID-19 (ref have not had COVID-19) | Have had COVID-19, suspected by self or doctor | 1.38 | 0.89 | 2.15 | 1.24 | 0.76 | 2.03 |
|  | Have had COVID-19, confirmed by test | 1.11 | 0.66 | 1.88 | 1.06 | 0.59 | 1.91 |
| Stressed in the past week about catching/becoming ill from COVID-19 (ref stressed) | Not stressed | 1.80 | 1.19 | 2.72 | 1.91 | 1.19 | 3.07 |
| Initial COVID-19 vaccine intention (ref willing) | Initially uncertain about accepting a COVID-19 vaccine | 4.92 | 2.98 | 8.11 | 5.29 | 3.07 | 9.09 |
|  | Initially unwilling to accept a COVID-19 vaccine | 6.40 | 3.94 | 10.41 | 11.29 | 6.79 | 18.78 |

Note. 'Very likely' to accept a COVID-19 booster vaccine was the reference group in the multinomial regression model. Ethnic minority groups refers to Black, Asian and minority ethnicity. GCSE refers to General Certificate of Secondary Education. Data were weighted to the proportions of gender, age, ethnicity, country, and education obtained from the Office for National Statistics.

**Table S7.** Sensitivity analysis: Socio-demographic, COVID-19 related, and initial COVID-19 vaccine intent predictors of uncertainty and unwillingness to receive a COVID-19 booster vaccine using a multivariable multinomial regression including only participants who had had exactly two doses of a COVID-19 vaccine at follow-up (weighted, *N* = 12,297)

|  |  | Uncertain |  |  | Unwilling |  |  |
| --- | --- | --- | --- | --- | --- | --- | --- |
|  |  | RRR | 95% CI |  | RRR | 95% CI |  |
| Gender (ref male) | Female | 1.12 | 0.76 | 1.63 | 1.04 | 0.67 | 1.60 |
| Age (ref 60+) | 45-59 | 0.82 | 0.50 | 1.34 | 1.06 | 0.59 | 1.91 |
|  | 30-44 | 1.69 | 0.98 | 2.91 | 1.29 | 0.62 | 2.66 |
|  | 18-29 | 1.85 | 0.97 | 3.50 | 2.69 | 1.15 | 6.30 |
| Ethnicity (ref white) | Ethnic minority groups | 0.85 | 0.44 | 1.64 | 1.15 | 0.57 | 2.31 |
| Education (ref undergraduate degree or higher) | A-levels or vocational | 1.24 | 0.77 | 1.99 | 1.70 | 0.98 | 2.95 |
|  | Up to GCSE | 1.37 | 0.86 | 2.20 | 1.83 | 1.00 | 3.34 |
| Income (ref >£90,000) | £60,000-89,999 | 0.55 | 0.23 | 1.35 | 0.34 | 0.11 | 1.08 |
|  | £30,000-59,999 | 2.49 | 1.08 | 5.75 | 0.71 | 0.28 | 1.79 |
|  | £16,000-29,999 | 2.20 | 0.86 | 5.67 | 1.50 | 0.55 | 4.06 |
|  | <£16,000 | 2.91 | 1.06 | 7.97 | 1.95 | 0.71 | 5.33 |
| Employment status (ref not employed) | Employed | 3.12 | 1.92 | 5.06 | 1.49 | 0.85 | 2.62 |
| Living arrangement (ref alone) | Living with others, not including children | 0.77 | 0.43 | 1.38 | 0.83 | 0.41 | 1.68 |
|  | Living with others, including children | 1.69 | 0.89 | 3.23 | 1.77 | 0.86 | 3.61 |
| Area of dwelling (ref urban) | Rural | 0.85 | 0.53 | 1.34 | 1.03 | 0.54 | 1.94 |
| Smoking status (ref former or non-smoker) | Current smoker | 1.95 | 1.18 | 3.22 | 1.43 | 0.73 | 2.79 |
| Key worker status (ref not a key worker) | Key worker | 0.91 | 0.58 | 1.41 | 1.45 | 0.80 | 2.63 |
| Long-term physical health condition (ref condition) | No long-term physical health condition | 1.07 | 0.71 | 1.60 | 0.90 | 0.57 | 1.41 |
| Long-term mental health condition (ref condition) | No long-term mental health condition | 1.39 | 0.88 | 2.21 | 1.49 | 0.81 | 2.74 |
| Confidence in government to handle pandemic (ref high) | Low confidence in government to handle pandemic | 1.13 | 0.76 | 1.69 | 0.88 | 0.52 | 1.50 |
| Confidence in health system to handle pandemic (ref high) | Low confidence in health system to handle pandemic | 1.33 | 0.90 | 1.97 | 0.85 | 0.53 | 1.37 |
| Knowledge of COVID 19 (ref high) | Low knowledge of COVID-19 | 1.73 | 1.15 | 2.61 | 1.14 | 0.65 | 2.01 |
| Compliance across strict restriction periods (ref high) | Consistently low compliance with COVID-19 guidelines | 1.50 | 1.03 | 2.20 | 2.43 | 1.61 | 3.66 |
| Have had COVID-19 (ref have not had COVID-19) | Have had COVID-19, suspected by self or doctor | 1.23 | 0.78 | 1.94 | 1.12 | 0.67 | 1.87 |
|  | Have had COVID-19, confirmed by test | 1.07 | 0.63 | 1.83 | 1.16 | 0.63 | 2.12 |
| Stressed in the past week about catching/becoming ill from COVID-19 (ref stressed) | Not stressed | 1.90 | 1.24 | 2.91 | 1.83 | 1.11 | 3.00 |
| Initial COVID-19 vaccine intention (ref willing) | Initially uncertain about accepting a COVID-19 vaccine | 4.62 | 2.77 | 7.70 | 5.31 | 3.05 | 9.25 |
|  | Initially unwilling to accept a COVID-19 vaccine | 5.36 | 3.25 | 8.84 | 9.15 | 5.29 | 15.84 |

Note. 'Very likely' to accept a COVID-19 booster vaccine was the reference group in the multinomial regression model. Ethnic minority groups refers to Black, Asian and minority ethnicity. GCSE refers to General Certificate of Secondary Education. Data were weighted to the proportions of gender, age, ethnicity, country, and education obtained from the Office for National Statistics.

## References

1. Haas EJ, Angulo FJ, McLaughlin JM, Anis E, Singer SR, Khan F, et al. Impact and effectiveness of mRNA BNT162b2 vaccine against SARS-CoV-2 infections and COVID-19 cases, hospitalisations, and deaths following a nationwide vaccination campaign in Israel: an observational study using national surveillance data. The Lancet. 2021;397(10287):1819–29.

2. Mor V, Gutman R, Yang X, White EM, McConeghy KW, Feifer RA, et al. Short-term impact of nursing home SARS-CoV-2 vaccinations on new infections, hospitalizations, and deaths. J Am Geriatr Soc. 2021;

3. Falsey AR, Frenck Jr RW, Walsh EE, Kitchin N, Absalon J, Gurtman A, et al. SARS-CoV-2 neutralization with BNT162b2 vaccine dose 3. N Engl J Med. 2021;385(17):1627–9.

4. Levine-Tiefenbrun M, Yelin I, Alapi H, Katz R, Herzel E, Kuint J, et al. Viral loads of Delta-variant SARS-CoV2 breakthrough infections following vaccination and booster with the BNT162b2 vaccine. medRxiv. 2021;

5. Abu-Raddad LJ, Chemaitelly H, Butt AA. Effectiveness of the BNT162b2 Covid-19 Vaccine against the B. 1.1. 7 and B. 1.351 Variants. N Engl J Med. 2021;

6. Lopez Bernal J, Andrews N, Gower C, Gallagher E, Simmons R, Thelwall S, et al. Effectiveness of Covid-19 vaccines against the B. 1.617. 2 (Delta) variant. N Engl J Med. 2021;585–94.

7. Milne G, Hames T, Scotton C, Gent N, Johnsen A, Anderson RM, et al. Does infection with or vaccination against SARS-CoV-2 lead to lasting immunity? Lancet Respir Med. 2021 Dec 1;9(12):1450–66.

8. Schaefer GO, Leland RJ, Emanuel EJ. Making Vaccines Available to Other Countries Before Offering Domestic Booster Vaccinations. JAMA. 2021;326(10):903–4.

9. UK Government. Vaccinations in the UK | Coronavirus in the UK [Internet]. 2021 [cited 2021 Dec 6]. Available from: https://coronavirus.data.gov.uk/details/vaccinations

10. Office for National Statistics. Coronavirus (COVID-19) Infection Survey technical article: Analysis of characteristics associated with vaccination uptake [Internet]. 2021. Available from: https://www.ons.gov.uk/peoplepopulationandcommunity/healthandsocialcare/conditionsanddiseases/articles/coronaviruscovid19infectionsurveytechnicalarticleanalysisofcharacteristicsassociatedwithvaccinationuptake/2021-11-15

11. Paul E, Steptoe A, Fancourt D. Attitudes towards vaccines and intention to vaccinate against COVID-19: Implications for public health communications. Lancet Reg Health - Eur [Internet]. 2021 Feb 1 [cited 2021 Feb 22];1(100012). Available from: https://www.thelancet.com/journals/lanepe/article/PIIS2666-7762(20)30012-0/abstract

12. Rhodes A, Hoq M, Measey M-A, Danchin M. Intention to vaccinate against COVID-19 in Australia. Lancet Infect Dis. 2020 Sep;S1473309920307246.

13. Robertson E, Reeve KS, Niedzwiedz CL, Moore J, Blake M, Green M, et al. Predictors of COVID-19 vaccine hesitancy in the UK household longitudinal study. Brain Behav Immun. 2021 May 1;94:41–50.

14. Sherman SM, Smith LE, Sim J, Amlôt R, Cutts M, Dasch H, et al. COVID-19 vaccination intention in the UK: results from the COVID-19 vaccination acceptability study (CoVAccS), a nationally representative cross-sectional survey. Hum Vaccines Immunother. 2021;17(6):1612–21.

15. Cascini F, Pantovic A, Al-Ajlouni Y, Failla G, Ricciardi W. Attitudes, acceptance and hesitancy among the general population worldwide to receive the COVID-19 vaccines and their contributing factors: A systematic review. EClinicalMedicine. 2021;40:101113.

16. Stead M, Jessop C, Angus K, Bedford H, Ussher M, Ford A, et al. National survey of attitudes towards and intentions to vaccinate against COVID-19: implications for communications. BMJ Open. 2021;11(10):e055085.

17. Loomba S, de Figueiredo A, Piatek SJ, de Graaf K, Larson HJ. Measuring the impact of COVID-19 vaccine misinformation on vaccination intent in the UK and USA. Nat Hum Behav. 2021;5(3):337–48.

18. Office for National Statistics. Coronavirus and the social impacts on Great Britain-3 December 2021 [Internet]. 2021 [cited 2021 Dec 6]. Available from: https://www.ons.gov.uk/peoplepopulationandcommunity/healthandsocialcare/healthandwellbeing/bulletins/coronavirusandthesocialimpactsongreatbritain/latest

19. Office for National Statistics. Coronavirus and the social impacts on Great Britain: 17 December 2021 [Internet]. 2021 [cited 2022 Jan 7]. Available from: https://www.ons.gov.uk/peoplepopulationandcommunity/healthandsocialcare/healthandwellbeing/bulletins/coronavirusandthesocialimpactsongreatbritain/17december2021

20. Office for National Statistics. Coronavirus and changing attitudes towards vaccination, England [Internet]. 2021 [cited 2021 Dec 9]. Available from: https://www.ons.gov.uk/peoplepopulationandcommunity/healthandsocialcare/healthandwellbeing/bulletins/coronavirusandchangingattitudestowardsvaccinationengland/7to16september2021#attitudes-towards-covid-19-booster-vaccines

21. Office for National Statistics. Population estimates for the UK, England and Wales, Scotland and Northern Ireland [Internet]. 2020 May [cited 2020 Sep 30]. Available from: https://www.ons.gov.uk/peoplepopulationandcommunity/populationandmigration/populationestimates/bulletins/annualmidyearpopulationestimates/mid2018

22. Hainmueller J, Xu Y. Ebalance: A Stata package for entropy balancing. J Stat Softw. 2013;54(7).

23. StataCorp. Stata Statistical Software: Release 16. College Station, TX: StataCorp LP; 2019.

24. Lazarus JV, Ratzan SC, Palayew A, Gostin LO, Larson HJ, Rabin K, et al. A global survey of potential acceptance of a COVID-19 vaccine. Nat Med. 2020 Oct 20;

25. de Figueiredo A, Simas C, Karafillakis E, Paterson P, Larson HJ. Mapping global trends in vaccine confidence and investigating barriers to vaccine uptake: a large-scale retrospective temporal modelling study. The Lancet. 2020;396(10255):898–908.

26. Dyer O. Covid-19: Rich countries’ booster plans will impede global vaccination, experts say. BMJ. 2021;374:n2353.

27. Institute of Global Health Innovation. UK and USA attitudes towards COVID-19 booster vaccines [Internet]. Imperial College London; 2021. Available from: https://www.imperial.ac.uk/media/imperial-college/institute-of-global-health-innovation/UK_US-vaccine-insights_ICL-YouGov-Covid-19-Behaviour-Tracker_20210625_final.pdf

28. Czeisler MÉ, Rajaratnam SM, Howard ME, Czeisler CA. COVID-19 Vaccine Intentions in the United States—December 2020 to March 2021. MedRxiv. 2021;2021.05.16.21257290.

29. Abedi V, Olulana O, Avula V, Chaudhary D, Khan A, Shahjouei S, et al. Racial, Economic, and Health Inequality and COVID-19 Infection in the United States. J Racial Ethn Health Disparities. 2021;8(3):732–42.

30. Krieger N, Waterman PD, Chen JT. COVID-19 and Overall Mortality Inequities in the Surge in Death Rates by Zip Code Characteristics: Massachusetts, January 1 to May 19, 2020. Am J Public Health. 2020;110(12):1850–2.

31. Tan AX, Hinman JA, Abdel Magid HS, Nelson LM, Odden MC. Association Between Income Inequality and County-Level COVID-19 Cases and Deaths in the US. JAMA Netw Open. 2021 May 3;4(5):e218799.

32. Whittle RS, Diaz-Artiles A. An ecological study of socioeconomic predictors in detection of COVID-19 cases across neighborhoods in New York City. BMC Med. 2020 Sep 4;18(1):271.

33. Niño M, Harris C, Drawve G, Fitzpatrick KM. Race and ethnicity, gender, and age on perceived threats and fear of COVID-19: Evidence from two national data sources. SSM -Popul Health. 2021;13:100717.

34. Wright L, Fancourt D. Do predictors of adherence to pandemic guidelines change over time? A panel study of 22,000 UK adults during the COVID-19 pandemic. Prev Med. 2021 Dec 1;153:106713.

35. Chu DK, Akl EA, Duda S, Solo K, Yaacoub S, Schünemann HJ, et al. Physical distancing, face masks, and eye protection to prevent person-to-person transmission of SARS-CoV-2 and COVID-19: a systematic review and meta-analysis. The Lancet. 2020;395(10242):1973–87.

36. Stead M, Jessop C, Angus K, Bedford H, Ussher M, Ford A, et al. National survey of attitudes towards and intentions to vaccinate against COVID-19: implications for communications. BMJ Open. 2021;11(10):e055085.

37. World Health Organization. Best practice guidance: How to respond to vocal vaccine deniers in public. 2016.

